# The Long-Term Outcomes of Neural Tube Defects in Eastern Africa: A Systematic Review and Meta-Analysis

**DOI:** 10.64898/2026.06.26.26356687

**Authors:** Hope M Mwangudzah, Josiline Chemutai, Belinda J Njiro, Rosie Cornish, Sarah J Lewis, Grace M Power

**Author notes:** Correspondence: Sarah J. Lewis, Department of Population Health Sciences, The Medical School, University of Bristol, Augustine’s Courtyard, Orchard Lane, Bristol BS1 5DS, UK. These authors contributed equally (Rosie Cornish, Grace M Power).

## Abstract

**Background:** Neural tube defects (NTDs) are preventable congenital malformations that disproportionately affect low and middle-income countries and contribute to disability and mortality. Evidence on the long-term outcomes of children and adolescents with NTDs in Eastern Africa has not been comprehensively synthesised. We conducted a systematic review and meta-analysis to assess survival, complications, and functional outcomes among children with NTDs in this region.

**Methods:** We searched PubMed, MEDLINE (Ovid), Cochrane Library, Web of Science, and Africa Index Medicus from the earliest records to April 2025. Two reviewers independently screened studies, assessed quality, and extracted data. We included studies reporting outcomes beyond one year of age, except mortality, which was assessed from birth onwards. Random-effects meta-analyses were undertaken when at least two sufficiently studies were available; otherwise, findings were synthesised narratively.

**Results:** Of 597 articles screened, 16 studies involving 2,340 children with NTDs met the inclusion criteria. Pooled cumulative mortality was 23% (95% CI: 12–36%) in the neonatal period, 9% (95% CI: 2–33%) during infancy, 22% (95% CI: 16–28%) in toddlers, 37% (95% CI: 30–44%) in pre-school-aged children and 45% (95% CI: 36–54%) in school-aged children. The pooled prevalence of hydrocephalus was 41% (95% CI: 34–48%) with little variation by age. Neurogenic bladder increased from 53% (95% CI 49-81%) in toddlers to 83% (95 % CI 72-91%) in adolescents, while impaired mobility affected about 61% (95% CI 48-72%) of adolescents. School enrolment was 53% (95% CI: 39-67%), with 9% (95% CI: 4%–19%) in specialized education. Speech, hearing, and bowel dysfunction were understudied (≤ 2 studies each).

**Conclusion:** Many children with NTDs in the Eastern African region survive beyond infancy but frequently experience hydrocephalus, neurogenic bladder and motor impairment. Longitudinal studies, context-specific guidelines, and follow-up systems are urgently needed to improve care and long-term outcomes.

**PROSPERO registered number:** CRD420251019868

**What is already known on this topic:**

- NTDs carry a substantial burden of mortality and lifelong disability. Survival and long-term outcomes in high-income settings are well-documented, but evidence from the Eastern African region is limited and fragmented.
- Existing studies provide an incomplete picture of survival, complications, functional outcomes, and school participation beyond infancy

**What this study adds:**

- This review demonstrates that mortality among children with NTDs increases substantially throughout childhood.
- Hydrocephalus, neurogenic bladder, impaired mobility, and low school enrolment are common, while bowel, speech, and hearing outcomes remain poorly reported.

**How this study might affect research practise or policy:**

- These findings underscore the need for structured long-term follow-up and multidisciplinary care for children with NTDs encompassing rehabilitation, continence care, and inclusive education.
- Future research should prioritise prospective cohort designs with standardised outcome measurement to generate context-specific evidence

## Background

Neural tube defects (NTDs) are congenital malformations resulting from incomplete closure of the neural tube during embryonic development, usually within the first four weeks after conception. They are among the most common neurological anomalies in Africa [1, 2], with particularly high incidence in East Africa [3], a subregion of Eastern Africa (EA). Regional estimates suggest a prevalence of around 3 per 1,000 births, affecting more than 1,000 infants annually. Despite strong evidence that NTDs are preventable through folic acid (FA) supplementation and staple food fortification [4–6], they continue to contribute substantially to disability and mortality in low and middle-income countries (LMICs).

Reducing NTD incidence in LMICs remains challenging. High rates of unplanned pregnancy, low uptake of preconception FA supplementation, limited access to fortified foods, and late initiation of antenatal care all contribute to persistently high NTD rates [7–9]. In the EA region, just over half of countries have adopted mandatory FA fortification; however, implementation is often incomplete [10], coverage is uneven, and FA supplementation is frequently administered after the critical periconceptional window, reducing its effectiveness for prevention [11].

NTDs range from anencephaly, which is universally fatal, to spina bifida, which has a more variable prognosis. Myelomeningocele (MMC), the most common survivable form [12], is associated with substantial long-term morbidity, including motor impairment or paralysis, neurogenic bladder and bowel dysfunction, hydrocephalus and neurodevelopmental difficulties. These sequelae create a considerable burden of disability and lifelong healthcare needs, particularly in resource-constrained settings where specialised neurosurgical, rehabilitation and support services are limited.

Existing evidence syntheses have focused predominantly on NTD prevalence, incidence, prevention and early mortality, with comparatively little attention to long-term physical, neurological and psychosocial outcomes [13–15]. In High-Income Countries (HIC), a systematic review and several large cohort studies suggest that survival into late adolescence and early adulthood now exceeds 80-90% for individuals with spina bifida and around 70% for those with encephalocele among those with access to specialist care [16–19]. These gains are attributed to advances in neonatal intensive care, improved prenatal diagnosis, increasing use of termination for severe foetal anomalies and widespread mandatory FA fortification, which has lowered the incidence of high-level lesions associated with poorer survival [20, 21].

By contrast, long-term outcome data from LMICs, including the EA region, are scarce. Inadequate health information systems, incomplete mortality ascertainment and limited infrastructure for long-term follow-up mean that many children born with NTDs are not captured in routine data. In this context, clinical practice and policy in the EA region often default to evidence generated in HICs where disease patterns, availability of specialist services, and social support structures differ substantially. As a result, HIC-derived recommendations are only partially applicable and may misestimate service needs and miss opportunities to prevent complications through feasible context-appropriate interventions.

The 77^th^ World Health Assembly recently prioritized improving care for children with congenital anomalies in LMICs [22, 23], highlighting the need to strengthen diagnosis, treatment and long-term follow-up. To translate this global commitment into effective practice in the EA region, robust regional outcome data are needed to inform context-specific guidelines, service delivery models and investment decisions, and planning for neurosurgical, rehabilitation, and educational services. To date, no comprehensive synthesis of long-term outcomes among individuals with NTDs in the EA region, where the burden of NTDs is disproportionately high [24–26], and healthcare resources are particularly constrained, has previously been published.

We therefore conducted a systematic review and meta-analysis of long-term outcomes associated with NTDs in this region. Our objectives were to: i) estimate survival among individuals with NTD; ii) determine the prevalence of major complications, including hydrocephalus, neurogenic bladder, paralysis, bowel dysfunction, and neurodevelopmental delays; and iii) assess school enrolment rates, including special education. This review aimed to provide region-specific evidence to support adaptation of international recommendations to the realities of the EA region health systems.

## Methods

This systematic review followed the recommendations outlined in the Joanna Briggs Institute (JBI) Manual for Evidence Synthesis [27] and adhered to the PRISMA checklist [28], as detailed in the supplementary information (SI) |SI Text. The protocol is available in SI Text 2.

### Patient and Public Involvement

Patient and public involvement was not relevant for this systematic review, as it relied solely on analysis of previously published studies. No patients or members of the public were directly involved in the design, conduct, reporting, or dissemination of this research.

### Search strategy and eligibility criteria

We systematically searched for studies on long-term NTD outcomes among individuals under 18 years. We used a predefined search strategy (SI Text 3) to search PubMed, MEDLINE (Ovid), the Cochrane Library, Web of Science, and Africa Index Medicus for English-language articles from the earliest available records through April 2025. We also screened the reference lists of all eligible studies. We included original scientific articles reporting cross-sectional or cohort studies from any healthcare setting (e.g., primary care, hospitals, or outpatient clinics) and from the EA region (Rwanda, Mauritius, Mozambique, Zimbabwe, Malawi, Zambia, Mayotte, Kenya, Tanzania, Uganda, Burundi, Ethiopia, South Sudan, Somali Eriteria, Seychelles, Sudan, Comoros, Djibout, and Madagasca). Detailed inclusion and exclusion criteria are provided in (SI Text 4)

### Study Selection and Data Extraction

Search results were exported to the Covidence web platform for reference management [29], and duplicates were removed. Two reviewers (HMM and JC) independently screened titles and abstracts; potentially eligible studies identified at this stage were retrieved for full-text assessment. Inter-reviewer agreement was evaluated at both the title and abstract screening stage and the full-text review stage. To quantify concordance between reviewers, we calculated percentage agreement and Cohen’s kappa statistic. Kappa values were interpreted according to the Landis and Koch [30] benchmarks: 0.21–0.40 indicating fair agreement, 0.41–0.60 moderate, 0.61–0.80 substantial, and 0.81–1.00 almost perfect agreement. Any discrepancies at either stage were resolved through discussion and consensus with additional reviewers (BJN, SJL, RC, and GMP).

Two independent reviewers (HMM and JC) extracted data using a customised Covidence template. Extracted information included study title, region type (urban/rural), country, settings, study design, sample size, mean age at baseline and follow-up, distribution of biological sex, NTD subtype, and reported outcomes by age (including prevalence data on mortality, hydrocephalus, paralysis, urinary dysfunction, school attendance, neurodevelopmental delays, and bowel dysfunction). Data were cross-checked, and discrepancies were resolved by consensus.

### Risk of bias assessment

Five reviewers (HMM, JC, SJL, RC, and GMP) assessed risk of bias for each outcome in every study using the risk of bias in measuring prevalence studies of mental health (RoB-PrevMH) tool [31], with at least three reviewers independently assessing each study. The RoB-PrevMH tool was specifically developed to assess disease prevalence and evaluate selection bias (sample representativeness and response bias) and information bias (outcome measurement bias). Among the cohort studies, we assessed response bias based on the proportion of participants who were lost to follow-up, as this may be associated with outcomes in our review. We categorized response bias as high risk when attrition exceeded 30%, moderate risk when it was 10–30%, and low risk when it was below 10% [32, 33], consistent with thresholds commonly used by Cochrane authors. Each domain was then rated as having low, moderate, or high risk of bias.

When information was ambiguous, the domain was rated as unclear (SI Table 3a–3g), and disagreements were resolved through discussion.

### Data synthesis and statistical analysis

We provided narrative descriptions for all pre-specified outcomes: mortality, hydrocephalus, neurogenic bladder, paralysis (typically reported as mobility function impairment), neurodevelopmental delays, school attendance and bowel dysfunction. Where there were two or more sufficiently similar studies of the same outcome, we conducted a meta-analysis to estimate the pooled prevalence. We also conducted subgroup analyses by age group and NTD subtype, where possible for each outcome. Age was stratified into six developmental groups according to the Munich age classification system, [34], which accounts for physiological and anatomical differences across childhood: neonatal (0–27 days), infancy (28 days to 12 months), toddlerhood (13 months to <2 years), preschool (3–5 years), school age (6–11 years), and adolescence (12–17 years). Prevalence and 95% confidence intervals were calculated using the inverse variance method with a random-effects model, and the I^2^ statistic quantified study heterogeneity. All analyses were conducted in R software (version 4.5.3) [35] with the meta package [36]. To assess the robustness of the pooled estimates to attrition bias, we performed a sensitivity analysis excluding studies that reported a loss to follow-up greater than 30%. When subtypes were clearly identified, chi-square tests were used to compare outcomes across subtypes; Fisher’s exact test was used when cell counts were below five. The risk of publication bias was not formally assessed because fewer than ten studies contributed to each long-term outcome analysis, consistent with guidance that funnel plots and Egger’s test are unreliable with small study numbers [37, 38].

## Results

Our search identified 943 published records, of which 598 remained after removing duplicates. After excluding 491 articles during the title and abstract screening, 107 full-text manuscripts were assessed for eligibility. Sixteen articles met the inclusion criteria: five cross-sectional studies [39–43] and eleven prospective or retrospective cohort studies [44–54]. Inter-reviewer agreement was fair, 77% at the titles and abstracts screening stage, and substantial, 90% at full-text review. Cohen’s kappa was 0.34 (95% CI: 0.26 to 0.42) for titles and abstracts, and 0.69 (95% CI: 0.55 to 0.83) for full-text. The study, selection process, and final sample are summarised in the PRISMA flow chart (Fig. 1).

**Fig 1.**
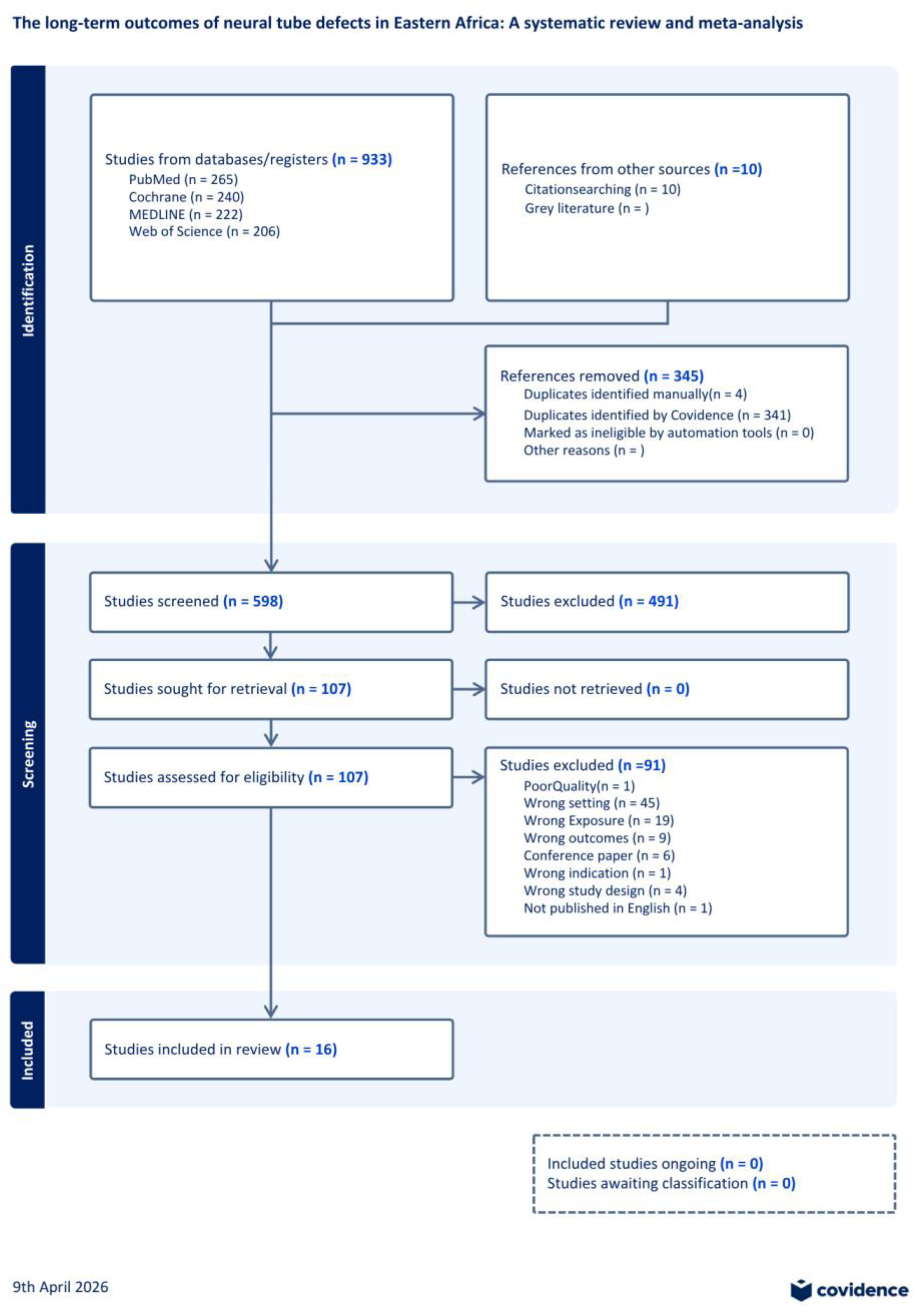
PRISMA flow chart. The flow chart illustrates the output from the article identification and selection process, namely: searching, title and abstract screening, and full-text screening for eligibility (both exclusion and inclusion)

### Characteristics of included studies

Studies were identified from four of the 20 United Nations (UN) defined EA countries [55] Ethiopia [39, 45, 50], Kenya [43, 46, 49], Tanzania [48, 51], and Uganda [40–42, 44, 47, 52–54]. Half (8/16) of the included studies were conducted in Uganda. Of note, three study cohorts from the Cure Children’s Hospital of Uganda (CCHU) were reported across seven publications: Bannik [40, 41], Sims-William [42, 44, 47, 53], and Benjamim [54]. Overall, 16 studies were included in this review, representing 12 unique study populations (Table 1). Among these publications, 14 examined MMC as a distinct entity [39–52], one examined meningocele [43], one examined spina bifida occulta [52], one examined closed NTDs [43], two examined encephalocele [52, 54], and one study investigated anencephaly [52].

**Table 1:**
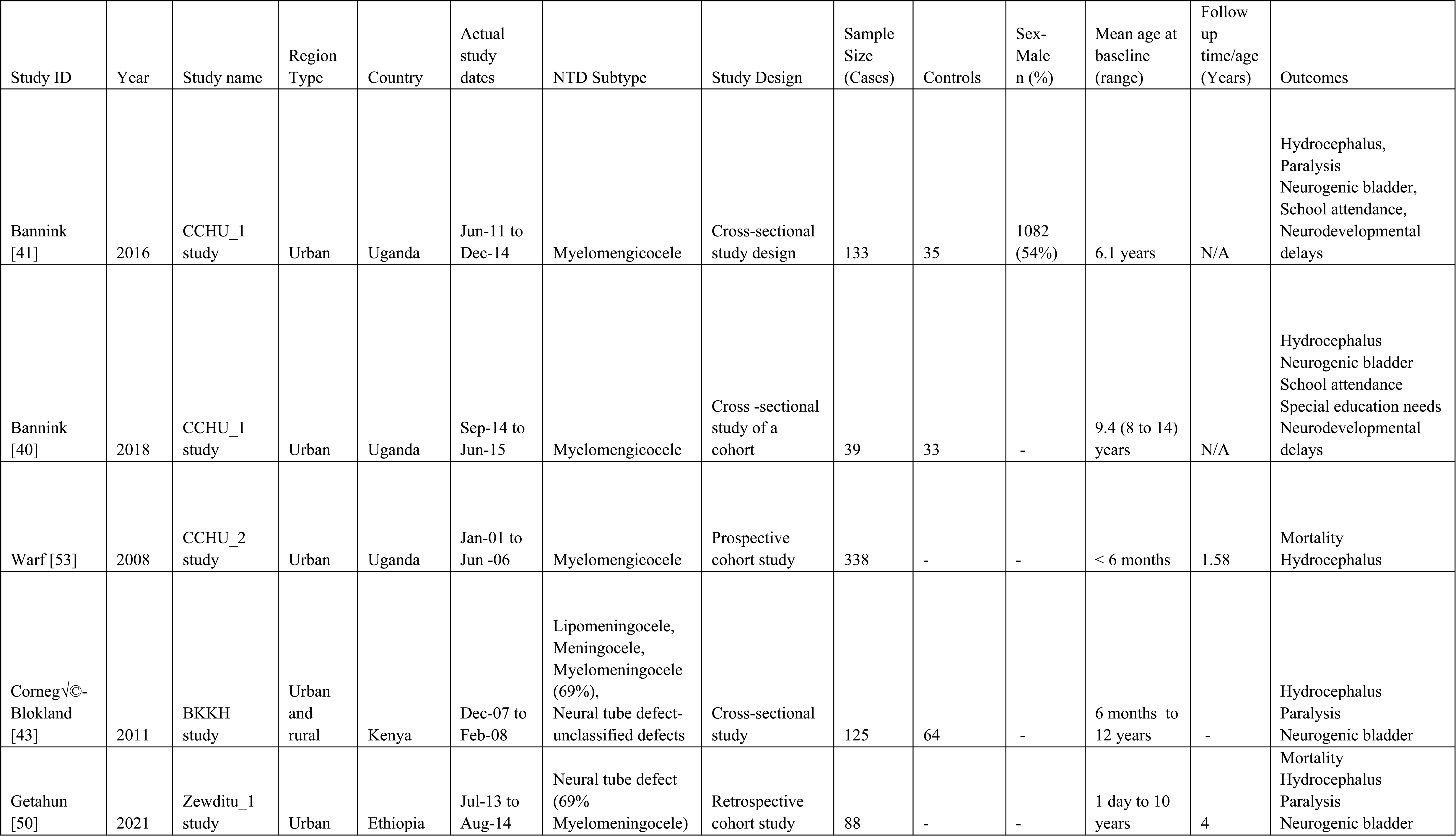
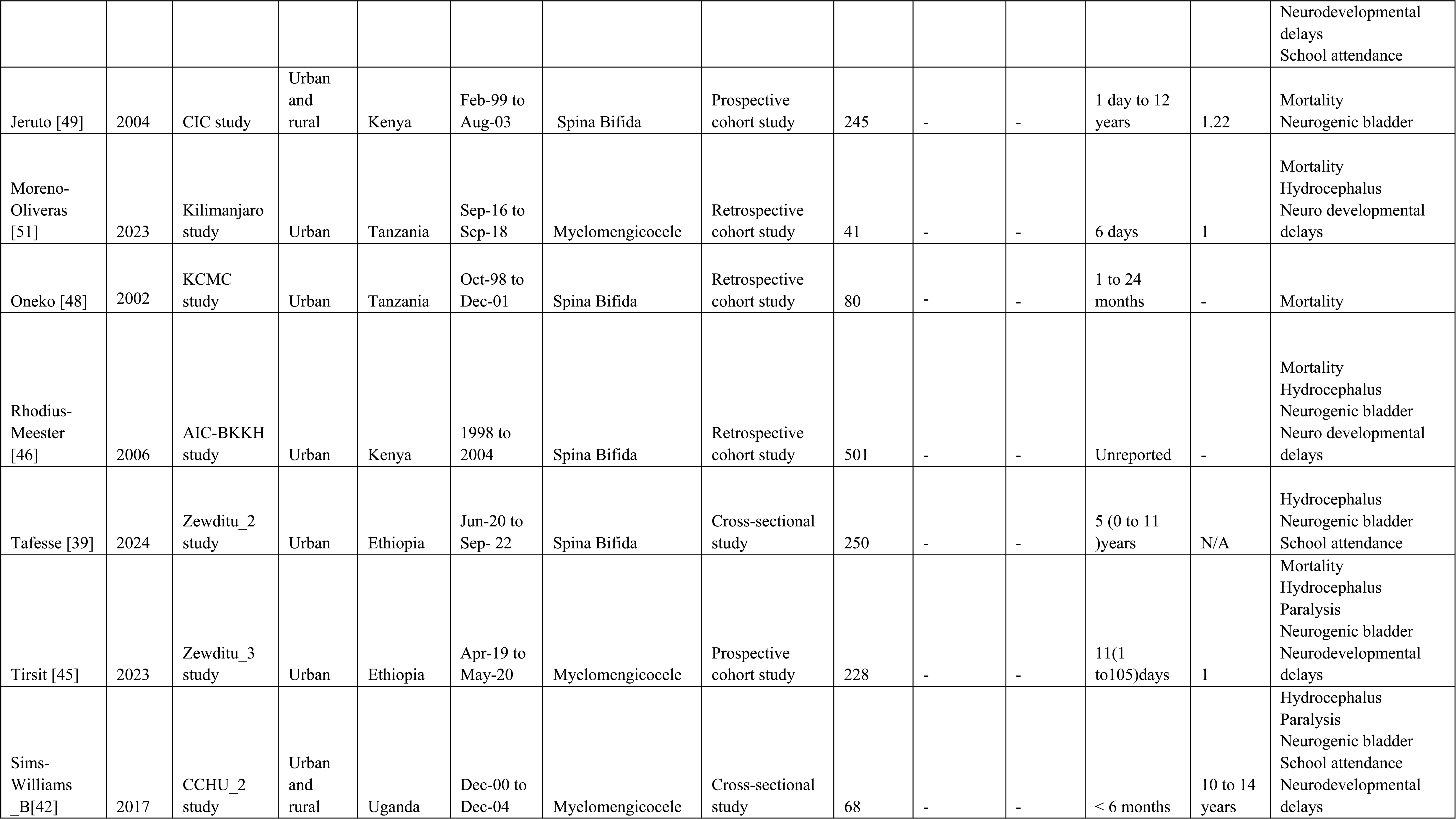
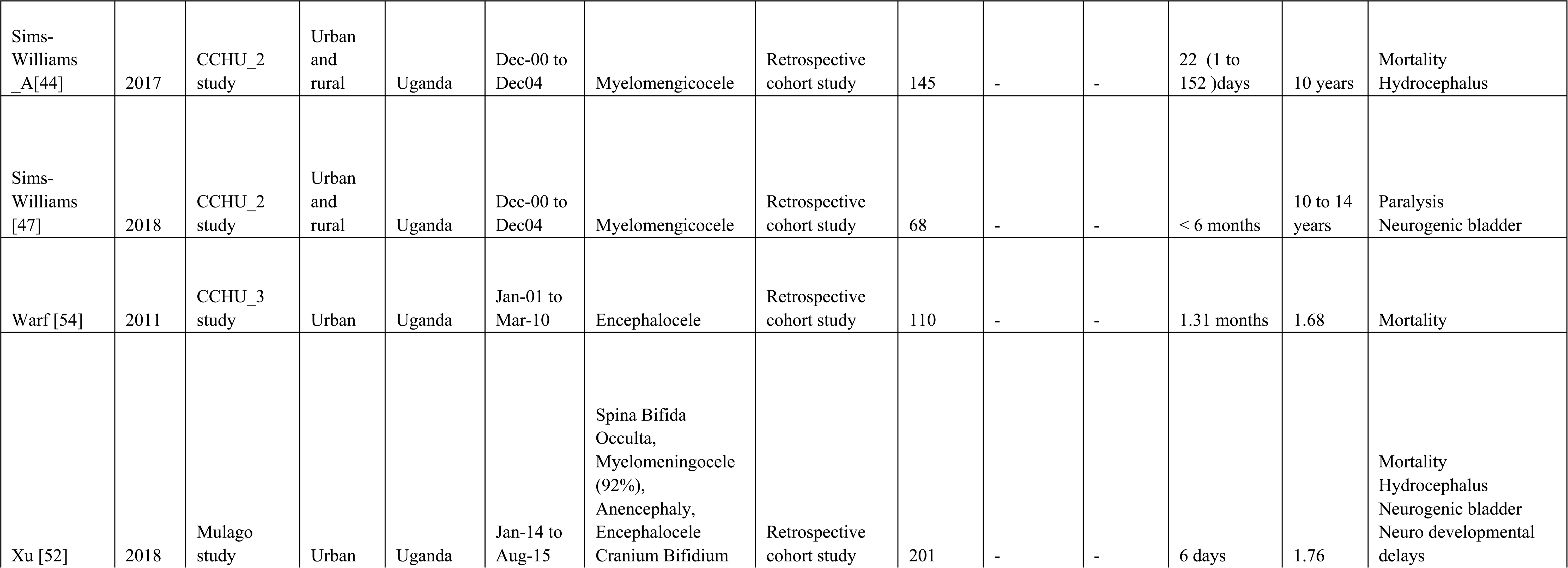
Study characteristics, CCHU (CURE Children’s Hospital Uganda), BKKH (Bethany Kids Kijabe Hospital), CIC (clean intermittent catheterization), KCMC (Kilimanjaro Christian Medical Centre), AIC-BKKH ( AIC - Bethany Kids Kijabe Hospital).

Sample sizes across included studies ranged from 41 to 501 participants, yielding a total of 2340 children with NTDs and 132 controls across 16 studies [39–54]. Among children with NTDs, ages ranged from birth to 14 years. In studies that reported the sex distribution, 1,082 of 2,008 children (54%) were male. Subtype analysis was constrained by insufficient details in the primary studies. Comparative statistics were feasible only for MMC and encephalocele subtypes and were limited to mortality [13 months–2 years] and hydrocephalus [all ages]. These two subtypes were included solely because of available reporting, not a predefined clinical hypothesis. Most other cases were grouped under broad non-specific labels such as spina bifida-unspecified, mixed spina bifida, or unclassified-NTDs.

### Mortality

Eight cohort studies (prospective and retrospective) reported mortality outcomes [44–46, 48, 50–52, 54]; three [44, 52, 54] contributed data at multiple time points. Pooled cumulative mortality estimates for each age group were 23% (95% CI: 12–36%) in the neonatal period, 9% (95% CI: 2–33%) during infancy, 22% (95% CI: 16–28%) in toddlers, 37% (95% CI: 30–44%) in pre-school-aged children and 45% (95% CI: 36–54%) in school-aged children (Fig. 2). One study [46], in which mortality data was not provided by age, was excluded from the age-group meta-analysis. Individual study estimates of mortality ranged from 3% to 45%. Heterogeneity was high in younger age groups (I² = 62–92%) and absent in preschool children (I² = 0%), but only two studies examined mortality among this age group.

**Figure 2:**
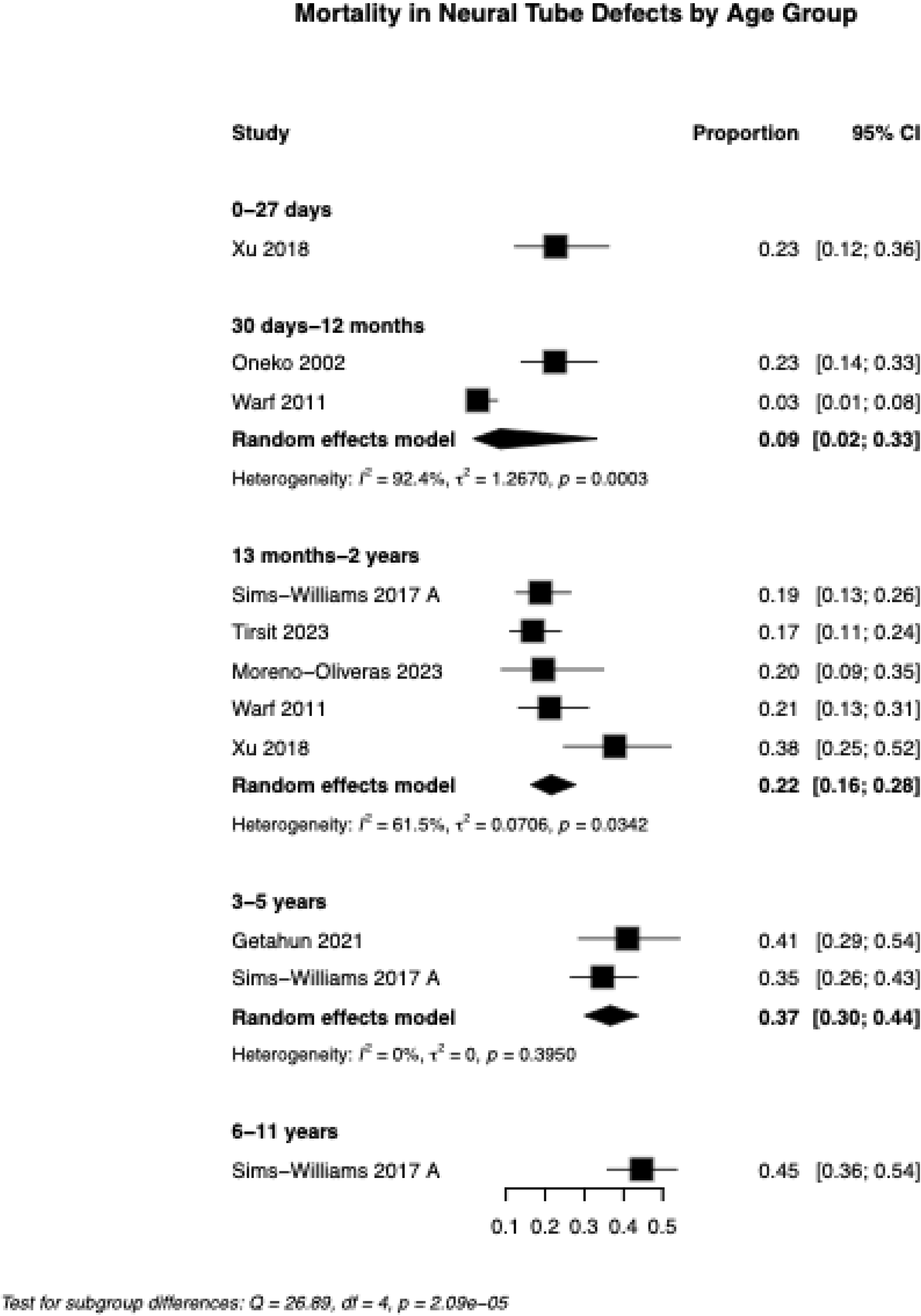
Forest plot of age-specific mortality in children with neural tube defects *Contributed data at multiple time points

Stratification by NTD subtype was restricted to the 13-month to 2-year age group to avoid including repeated measurements from the same participants across age categories, and because this was the age group with the most available studies. The subtype comparison (SI. Table 1) showed little evidence of differences (Q = 0.27, p = 0.60). Overall mortality was estimated at 18% (95% CI: 14–22%) in children with myelomeningocele (three studies, I² = 0%) and 21% (95% CI: 14–31%) in those with encephalocele (one study).

### Hydrocephalus

Data on Hydrocephalus (beyond one year of age) were available from three cross-sectional studies [39, 41, 43] and five cohort [44, 45, 51–53] studies. Across all age groups, the pooled prevalence was 41% (95% CI: 34-48%, I² = 76%). Individual study estimates ranged from 19% to 58% (Fig. 3). One study [46], which lacked age-stratified data, was excluded from the age-group meta-analysis. In a sensitivity analysis, two studies [45, 52] with > 30% attrition were excluded, and the pooled prevalence increased to 44% (95% CI:38-50%) (SI fig 2).

**Fig 3.**
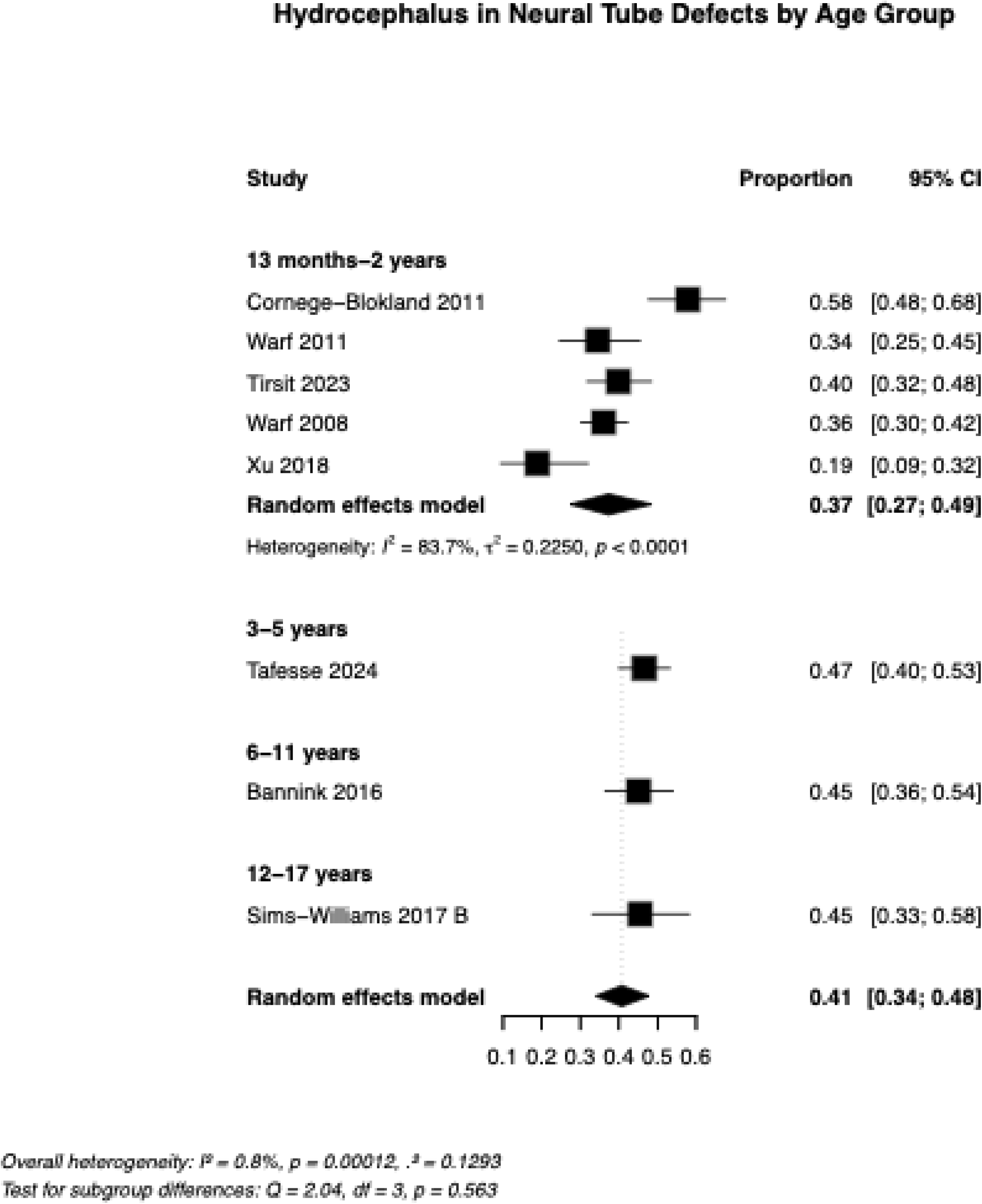
Forest plot of hydrocephalus prevalence in children with neural tube defects, pooled and stratified by age group

Stratification by NTD subtype (Q=0.36, p=0.55) showed no clear differences between subtypes. The pooled prevalence was 35% (95% CI: 25–45%) among children with encephalocele and 39% (95% CI: 34–43%) among those with MMC (3 studies, I² = 0%; SI Table 1).

### Neurogenic bladder

Long-term neurogenic bladder outcomes (beyond one year of age) were reported in four cross-sectional [39–41, 43] and five cohort studies [42, 45, 47, 49, 52]. Across these nine studies, the pooled prevalence was 67% (95% CI: 49-81%, I² = 95%). Prevalence increased with age (Fig. 4), from 53% |(95% CI: 25-80%) among children aged 13 months < 2 years to 83% (95 % CI: 72-91%) among those aged 12-17 years. Between-study heterogeneity was substantial (I² = 95%, p < 0.0001). In a sensitivity analysis, we excluded two studies [45, 49] with >30% attrition; the resulting pooled prevalence decreased to 64% (95% CI 40-81%) (SI fig 2).

**Fig. 4.**
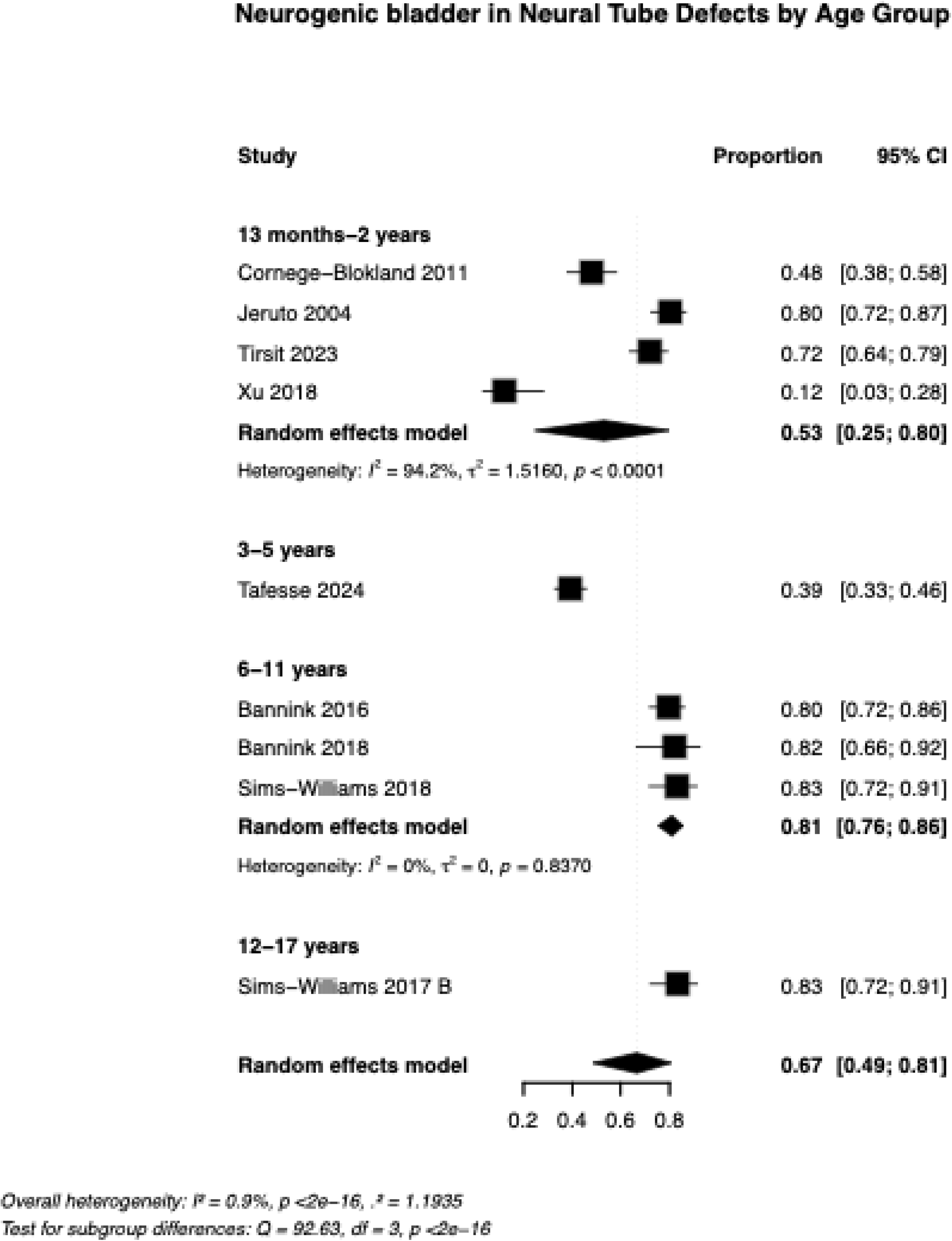
Forest plot of neurogenic bladder prevalence in children with neural tube defects, pooled and stratified by age group

### Paralysis

Paralysis (mobility function impairment) outcomes (beyond one year of age) were reported in two cross-sectional [41, 43] and four cohort [42, 45, 47, 50] studies. Overall, the pooled prevalence was 46% (95% CI: 39-54%, I² =71%, p=0.004). Prevalence was higher in school-age and adolescent groups than in toddlers and pre-schoolers (Fig. 5). Individual study estimates ranged from 31%-61%. In a sensitivity analysis, we excluded two studies [45, 50] with > 30% attrition; the resulting pooled prevalence increased to 50% (95 % CI: 42-59%) (SI fig 2).

**Fig. 5.**
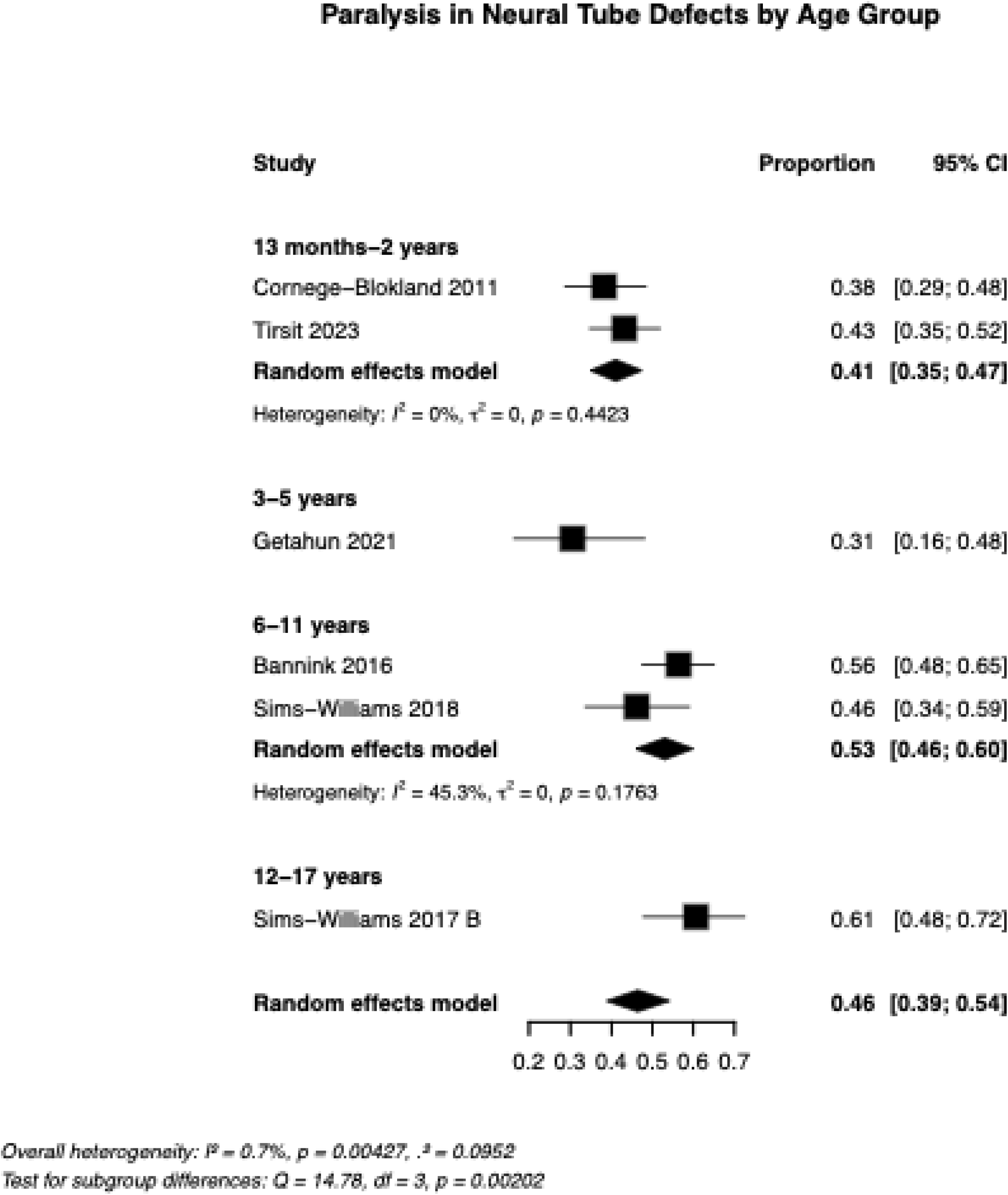
Forest plot of paralysis prevalence in children with neural tube defects, pooled and stratified by age group

### School attendance

Three cross-sectional [39–41] and two cohort studies [42, 50] reported school attendance, with a pooled prevalence of 53% (95% CI: 39-67%, I^2^=81%, p=0.003). Approximately half (47%) of all 3-5 year olds were attending school and this increased to 69% among 6-11 year olds but declined sharply in adolescence (36%) according to one study (Fig. 6). In one Ugandan study [42], only 9% (95% CI: 4%–19%) of children in school were enrolled in special education programs.

**Fig 6.**
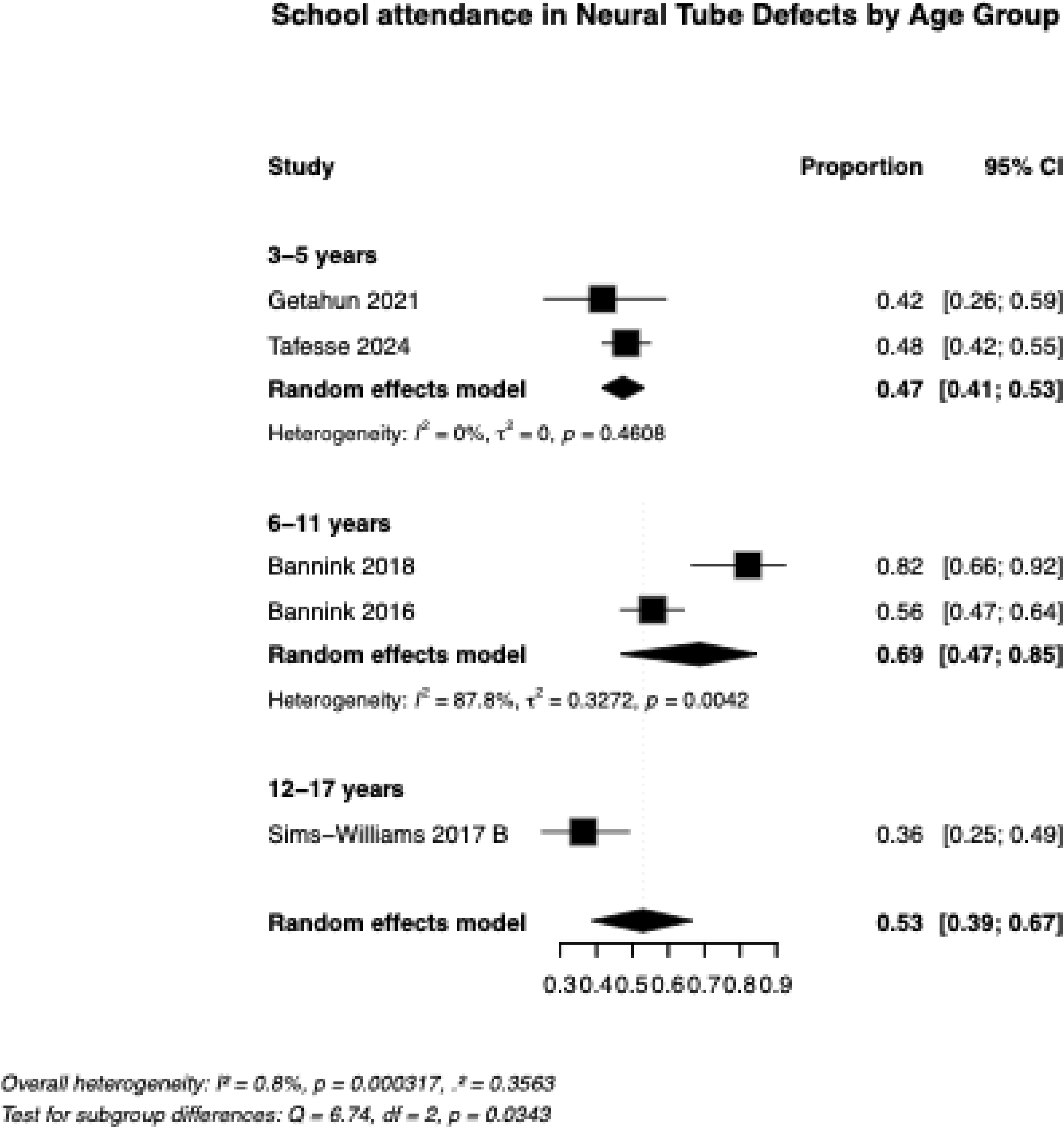
Forest plot of school attendance proportions in children with neural tube defects, pooled and stratified by age group

### Neurodevelopmental delays

Across one cross-sectional [40] and one cohort [52] study, the pooled prevalence of motor delay was 50% (95% CI: 41-60%) with low heterogeneity (I² = 27%, p = 0.26). The prevalence was 39% (95% CI: 23–58%, I² = 0%) in toddlers aged 13 months to 2 years, and 59% (95% CI: 42–74%) in school-age children aged 6-11 years (Fig. 7). Additional neurodevelopmental impairments were reported but not meta-analysed due to heterogeneous reporting. Two studies [41, 52] assessed cognitive delays using different formats (discrete vs continuous), preventing quantitative synthesis. One study [42] reported deafness in 6.1% (95% CI: 2-15%) of children, and another [52] reported speech impairment in 9.1% (95% CI: 3-24%).

**Fig. 7.**
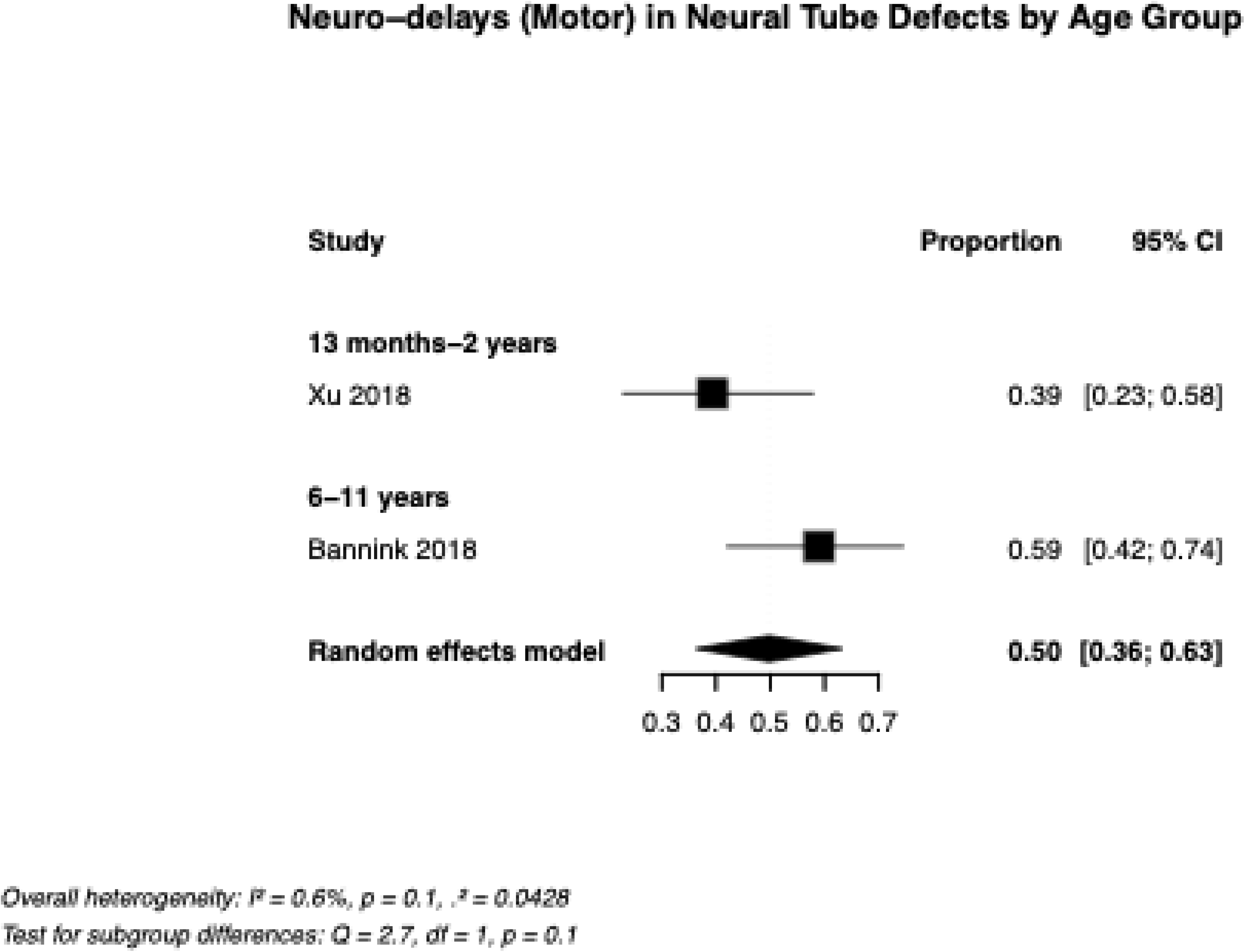
Forest plot of neurodevelopmental (motor) delays prevalence in children with neural tube defects, pooled and stratified by age group

### Bowel Dysfunction

Only one study reported bowel dysfunction among preschool children aged 3-5 years and reported a prevalence of 31% (95% CI: 18-47%).

### Risk of bias assessments

SI Tables 3a to 3g summarize the risk of bias assessment for all included studies. For mortality, the risk of selection bias was high in six of the nine studies [45, 46, 48, 50, 52, 54] because many could not account for deaths occurring before surgery. In contrast, two studies were rated as low risk, as they included all eligible participants [44, 51]. Similarly, response bias for mortality was high in five of the studies [45, 46, 48, 50, 52], with the remainder rated as low [44, 51], moderate [54], or unclear [49]. As mortality is an objective outcome, the risk of outcome measurement bias was rated as low across all studies.

For complications, sample selection methods varied. Ten studies [42, 44–47, 50–54] were judged to have a low risk of selection bias because they demonstrated inclusion of all eligible participants, clearly defined sampling frames, used consecutive recruitment, were conducted in a single national referral hospital, or, in one study, recruited from mobile satellite clinics across the country. One study was rated as high risk due to purposive sampling [41]. The remaining studies [39, 40, 43] were judged to have a moderate risk of selection bias owing to the exclusion of children with comorbidities, restrictive eligibility criteria, and non-random exclusions. Four studies were rated as low risk [44, 46, 47, 51], and several additional studies reported follow-up rates above 72 % [49, 53, 54]. Five cohort studies were judged to have a high risk of response bias [45, 46, 49, 50, 52]. In two of these studies, limited resources limited researchers’ ability to trace participants; moreover, nearly half of the participants had unknown status.

With respect to information bias, 88 % (14/16) of the studies [39–49, 51, 53, 54] collected outcome data via face-to-face interviews. The risk of bias for hydrocephalus was low in five of the 13 studies [45, 51–54] because they used objective measures, such as serial head circumference measurements or imaging-confirmed diagnoses. Three studies [40, 43, 50] relied on both parental reports and medical record extraction and were rated as having a moderate risk of bias. In the remaining five studies [39, 41, 42, 44, 46], the risk was unclear because the method used to establish the diagnosis was not specified. Information on neurogenic bladder was mainly obtained from parents or caregivers, who described continence status using an aided classification or the use of clean intermittent catheterization (CIC); these studies were rated as having a moderate risk of bias [39–43, 45, 47, 50, 52]. Only two studies used urodynamic testing combined with standardized clinical assessment. One was rated as low risk [49], whereas the other [47] was rated as moderate risk because the data collected during the study visit relied on retrospective review of notes.

Paralysis (mobility function impairment) was judged to have a low risk of bias in four of six studies [41–43, 47] that used standardized scales or neurological examinations, whereas the remaining two studies [45, 50] were rated as moderate risk because data were obtained from parents. School attendance was considered to have a moderate risk of bias [39–42, 50] in all five studies reporting this outcome, because attendance could not be validated against school records. Some neurodevelopmental outcomes [40–42] were rated as low risk of bias due to the use of validated instruments; however, other studies lacked such instruments and were therefore rated as having high [52] or unclear risk of bias [51]. The study evaluating bowel dysfunction [50] was judged to be at moderate risk of bias because it relied on subjective reporting.

## Discussion

This systematic review synthesises evidence on the long-term outcomes of neural tube defects (NTDs) in the EA region, where the disease burden is high, but data on outcomes are limited. The most consistently reported outcomes were mortality, hydrocephalus, neurogenic bladder, paralysis (mobility function impairment) and school attendance. In contrast, bowel function and neurodevelopmental outcomes were rarely assessed, highlighting important gaps in the regional evidence base. Direct comparison with other LMIC settings is severely constrained by the near-absence of published long-term outcome data from outside this region itself, a finding of significance that underscores both the novelty and necessity of this review.

Our findings suggest that although many children with NTDs in the EA region survive beyond infancy, substantial mortality continues throughout childhood. In the few cohorts with long-term follow-up, only about half were alive by late childhood [44, 50]. These figures are likely underestimated because many mortality studies had a high risk of attrition and did not include children who died before surgery. Although long -term survival data from LMICs remain scarce, comparison with data from HICs suggests a pronounced survival gap. Systematic reviews and large cohort studies from HICs generally report long-term survival rates of 85-90% for spina bifida into late adolescence or early adulthood, and around 70% for encephalocele among children with access to specialist care [16, 56–58]. Despite heterogeneity across studies, the contrast remains striking.

Several health systems and preventive factors likely contribute to this disparity. Access to timely neurosurgical intervention and intensive care, and structured long-term follow-up is far more limited locally in the EA region than in better-resourced settings [59], where dedicated NTD care pathways are often available. Prenatal diagnosis and, in some centres, fetal surgery can improve postnatal outcomes or result in termination of pregnancies affected by severe anomalies, meaning that HIC cohorts include relatively fewer of the most severe NTD cases [60, 61]. In addition, mandatory FA fortification in many HICs has substantially reduced NTD incidence [10] and is associated with a lower proportion of high-level lesions, which are linked to poorer survival. Our survival estimates may themselves underestimate the true mortality burden. Births and deaths are frequently under-reported in LMICs, and hospital-based cohorts capture only those infants who reach care [62, 63]. Children who die before presentation are unlikely to be represented in the available data. The combination of constrained access to specialised care and incomplete mortality ascertainment suggests that the true survival disadvantage may be greater than current estimates imply.

Hydrocephalus is the most commonly reported complication, which is consistent with the predominance of MMC and its strong association with Chiari II malformation, a condition that is universal among these children [64, 65]. Our pooled estimates suggest a substantial burden of hydrocephalus across childhood, although at lower levels than those reported in some high-resource and upper-middle-income country cohorts, where up to four in five children with MMC require cerebrospinal fluid diversion [66, 67]. Reports from other LMICs are broadly similar; however, these estimates are based on mixed paediatric populations and hospital-based studies [68]. Subtype analyses found no clear difference in hydrocephalus prevalence between MMC and encephalocele, although this comparison relied on limited evidence because only one encephalocele study was identified.

Several explanations may account for the lower prevalence we observed. Underdiagnosis is likely in settings with limited access to imaging, where hydrocephalus may only be recognised in its most advanced stages [69]. Variation in case definitions and the exclusion of unspecified NTD subtypes in some studies may also have contributed. In addition, higher early mortality among infants with severe hydrocephalus, combined with incomplete follow-up, could result in survival bias, whereby the children with the most severe disease are under-represented in older age groups. Without population-based surveillance using standardised diagnostic criteria, it remains difficult to distinguish true differences in hydrocephalus burden from artefacts of study design and health system capacity.

Neurogenic bladder was highly prevalent in older children and adolescents, and represents a major source of potentially preventable morbidity. In the few studies that used urodynamic testing and standardised criteria, neurogenic bladder was nearly universal among toddlers and adolescents with MMC, suggesting that underlying dysfunction is established early and persists throughout childhood [47, 49]. Lower prevalence estimates in other studies likely reflect less sensitive assessment methods, including reliance on caregiver reports.

Evidence from HICs indicates that early, proactive bladder management, especially CIC combined with anticholinergic therapy, is associated with 69% reduction in the odds of renal deterioration compared with delayed or reactive care [70, 71]. In our review, many centres reported using CIC [39–43, 45, 47, 49], but initiation was often delayed, and adherence was constrained, likely due to intermittent supply, absence of locally implemented standardised protocols, despite the availability of International Children’s Continence Society guidelines, and evidence synthesis on CIC in children with neurogenic bladder [72, 73]. Strengthening early identification of neurogenic bladder, ensuring consistent access to catheters and medications, and embedding standardised bladder care pathways could substantially reduce long-term renal and continence-related morbidity.

Longitudinal data indicate that motor deficits are established early in children with MMC, reflecting largely irreversible in utero neural tissue damage and limited spontaneous recovery [74–76]. Consistent with this, in one toddler study in our review, no child gained independent ambulation within one year of surgical repair [45]. The higher prevalence of motor impairment in adolescents compared with toddlers, although based on cross-sectional rather than longitudinal data, suggests possible deterioration over time due to tethered cord and progressive spinal deformity such as scoliosis [77–79].

Functional mobility remained severely limited at all ages; among studies reporting ambulation outcomes, only 8-27% of the children walked with assistive devices, and 22-47% were non-ambulatory [42, 45, 50]. Heterogeneous assessment methods constrain interpretation, as most studies used lesion-level classification rather than manual muscle testing, which more accurately predicts mobility [80–82]. These findings broadly mirror those from HICs, where 21-30% of children achieve functional ambulation after postnatal repair [83]. In HICs, prenatal repair yields substantially better ambulatory outcomes, with 50%-80%, achieving functional mobility [83–85], but such procedures are currently unavailable in the EA region settings represented in this review and are generally limited to a small number of highly specialised centres worldwide. Taken together, this pattern suggests that lesion level and the underlying neurological insults are major determinants of motor functions. Differences in perioperative care and rehabilitation are more likely to influence secondary complications and participation than to fundamentally alter baseline motor prognosis, in contrast to neurogenic bladder, where early proactive management can help prevent deterioration and reduce long-term morbidity.

School enrolment in the EA region is highly variable [65]. Children with NTDs face additional educational barriers, including incontinence [86] and mobility limitations [87], which further restrict school participation [88, 89]. In studies included in this review, enrolment was highest in children aged 6-11years and declined sharply in adolescence. Overall school enrolment, including special educational needs (SEN) programmes [42], was substantially lower than reported in HICs; for example, in Northern Ireland, 89% of children attend school, and in England, 78.6% of children with spina bifida receive SEN provision [90, 91]. In one Ugandan cohort, children with NTDs who were attending school showed better cognitive outcomes than those not in school, although their performance remained below that of typically developing siblings [41]. By contrast, data from Northern Ireland showed no cognitive differences among children born with spina bifida compared to peers without spina bifida, except in working memory [92], underscoring the value of sustained educational support. Working memory deficits are particularly important given their central role in academic learning [93]. Hydrocephalus adds further challenges, including visual attention deficits [94, 95], and school-related issues such as additional support needs, medical absences, and academic disengagement [96], highlighting the need for multi-level, inclusive interventions.

Speech and hearing outcomes remain under-researched. Evidence from a prenatal spina bifida surgery study in HIC showed no hearing impairments [84], yet 12% of children required speech therapy, indicating communication difficulties despite normal hearing. These difficulties may reflect motor speech control issues, cognitive-linguistic factors, or hydrocephalus/Chiari-related factors. In our review, only two of 16 studies [42, 52] assessed communication outcomes, highlighting this gap.

Neurogenic bowel dysfunction is similarly under-studied, mirroring the gaps in speech and hearing research. Only one of 16 studies reported on bowel dysfunction [50] and did not describe any structured management protocol. By contrast, in clinical practice in high-resourced settings, centres typically follow individualized, stepwise bowel programmes beginning with conservative measures and, when necessary, progressing to antegrade colonic enema (ACE) surgery [97, 98]. Given the major impact of bowel dysfunction and quality of life, there’s a clear need for prospective comparative research on standardized management pathways and patient-reported outcomes.

## Strengths

This systematic review has several strengths. It is, to our knowledge, the first comprehensive synthesis of long-term outcomes among individuals with NTDs in the EA region, where disease burden is high and long-term data have been limited. Unlike prior reviews focused on incidence, prevention, or short-term survival, it examines physical, neurological, functional, and neurodevelopmental outcomes across childhood and adolescence, providing a more complete picture of long-term morbidity. By emphasising outcomes beyond one year of age, it more clearly characterises survivorship and age-related changes in key complications. The use of systematic methods, predefined age strata and, where data allowed, subtype-specific analyses for mortality and hydrocephalus helps to address important gaps in the existing literature and offers region-specific evidence to inform service planning.

## Limitations

This review has several limitations, some of which arise from the underlying evidence. First, outcome definitions, measurement methods, and reporting varied substantially across studies, which restricted meta-analysis and required narrative synthesis for some outcomes. This heterogeneity also limited our ability to generate summary estimates and constrained subtype analyses to mortality and hydrocephalus, where reporting was more complete. Second, all included studies were of variable quality, with moderate or high risk of bias in outcome measurement and reporting. Although four studies had >30% attrition, sensitivity analyses suggested minimal impact on pooled prevalence estimates; nonetheless, loss to follow-up may have biased long-term outcomes towards children with better access to care. Third, clinically important outcomes such as bowel dysfunction, speech and hearing, and detailed cognitive function were rarely assessed, limiting our capacity to characterise the full spectrum of long-term morbidity. Fourth, age-stratified analyses were based on cross-sectional data rather than true longitudinal follow-up, so apparent age trends may partly reflect cohort differences rather than individual trajectories. Finally, restricting the search to English-language publications and not formally assessing publication bias may have led to over-representation of studies with positive or more complete findings, and underscores the need for more robust, prospective data from this region.

## Conclusion

In the Eastern African region, many children with NTDs now survive beyond infancy but frequently develop hydrocephalus, neurogenic bladder and severe motor impairment. School attendance also appears to decline during adolescence. Our review further shows that bowel, speech, and hearing outcomes are rarely measured, leaving important aspects of long-term morbidity poorly characterized. Improving outcomes will require earlier access to neurosurgical and urological care, and simple standardised protocols for hydrocephalus and bladder management that are feasible in this region. Longitudinal region-specific studies that include educational and patient-reported outcomes are essential to guide realistic planning of neurosurgical, rehabilitation, and inclusive education services for children with NTDs in the EA region.

## Data Availability

All data produced in the present work are contained in the manuscript

## Author contributions

**HMM:** Conceptualization, Methodology, Investigation, Data Curation, Validation, Visualization, Project Administration, Writing – Original Draft, Writing – Review & Editing; **JC:** Investigation, Data Curation, Validation; **BJN:** Investigation, Data Curation, Validation; **RC:** Conceptualization, Methodology, Supervision, Data Curation, Validation, Writing – Review & Editing; **SJL:** Conceptualization, Methodology, Supervision, Data Curation, Validation, Writing – Review & Editing; **GMP:** Conceptualization, Methodology, Supervision, Data Curation, Validation, Writing – Original Draft, Writing – Review & Editing.

---

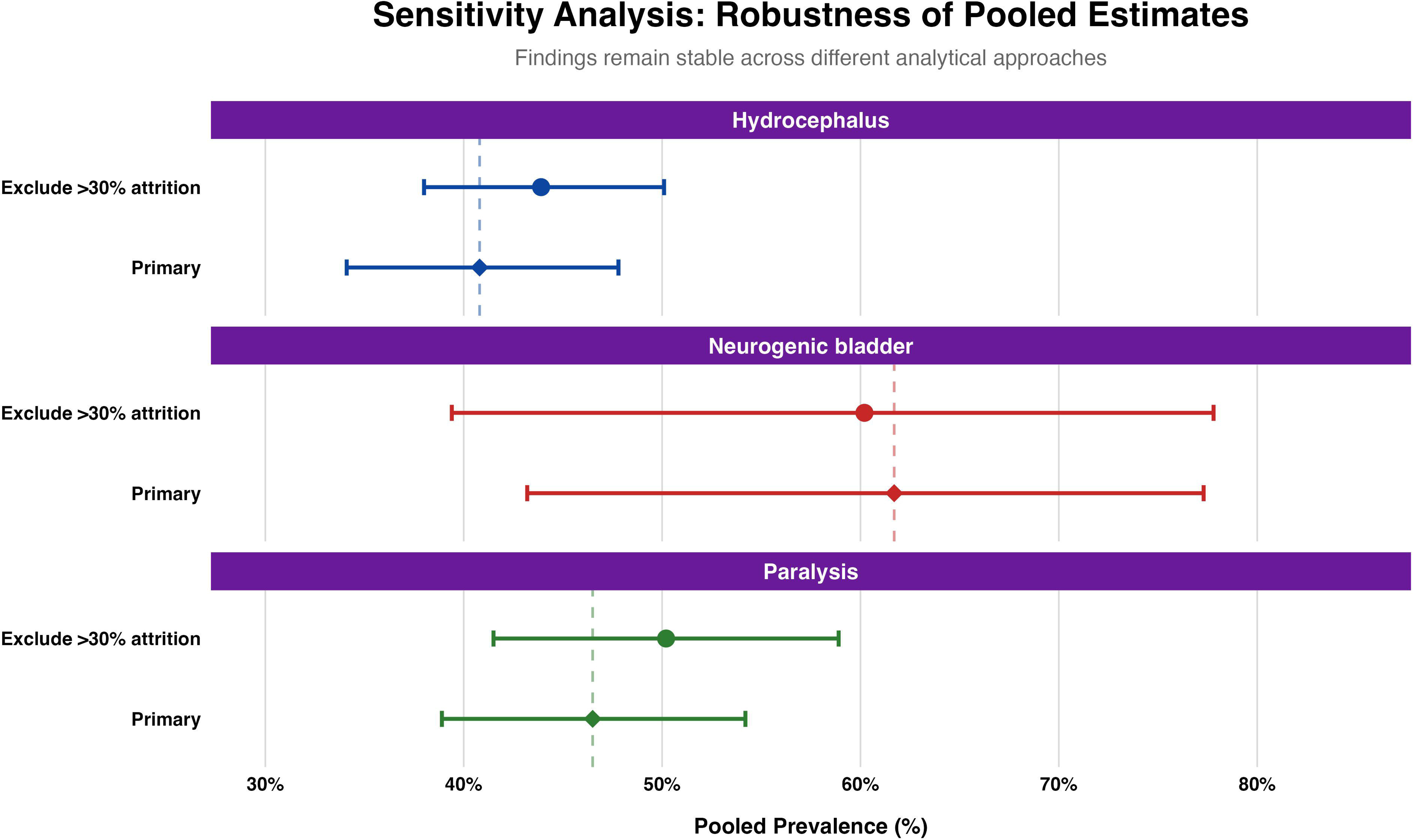

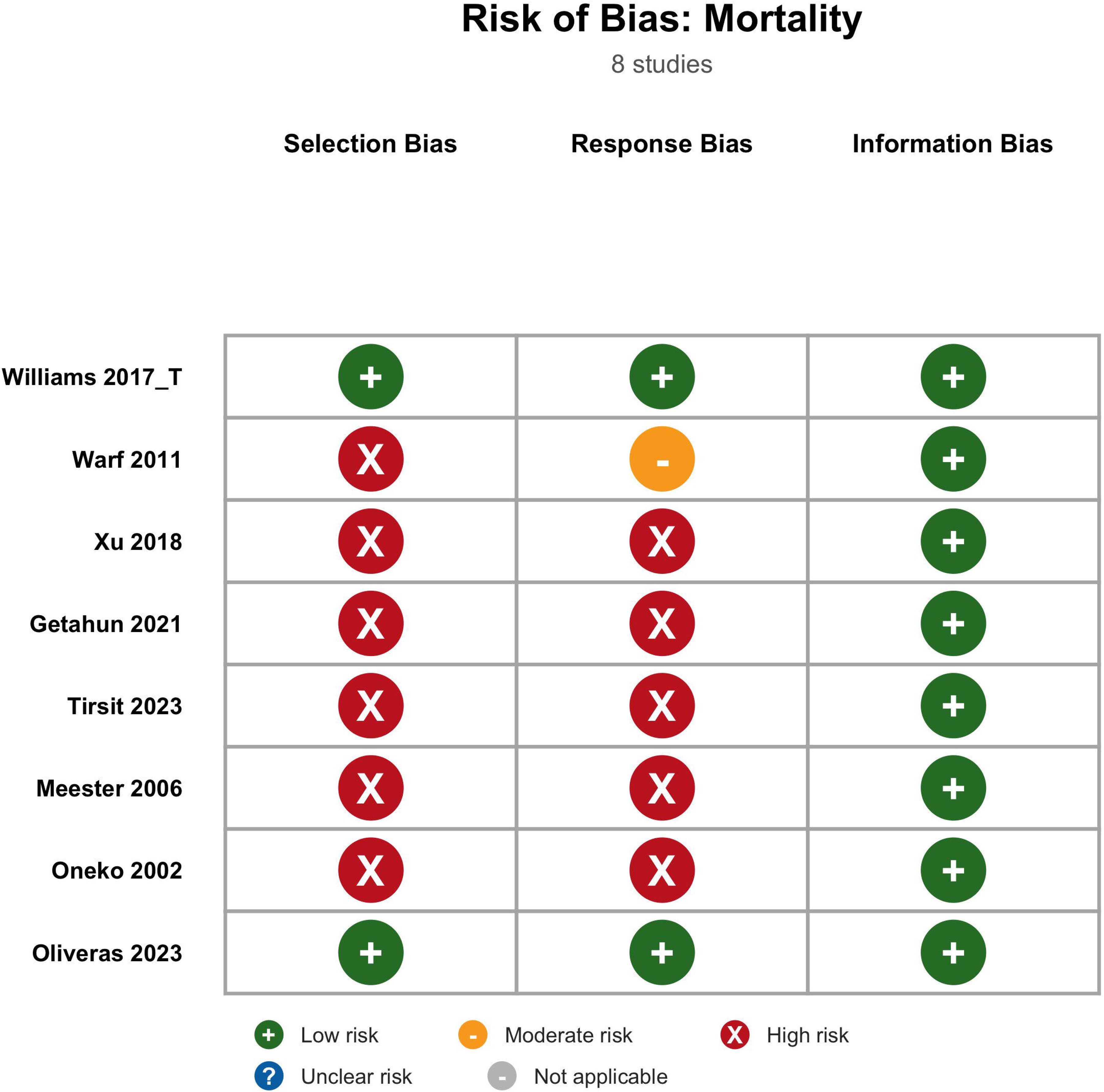

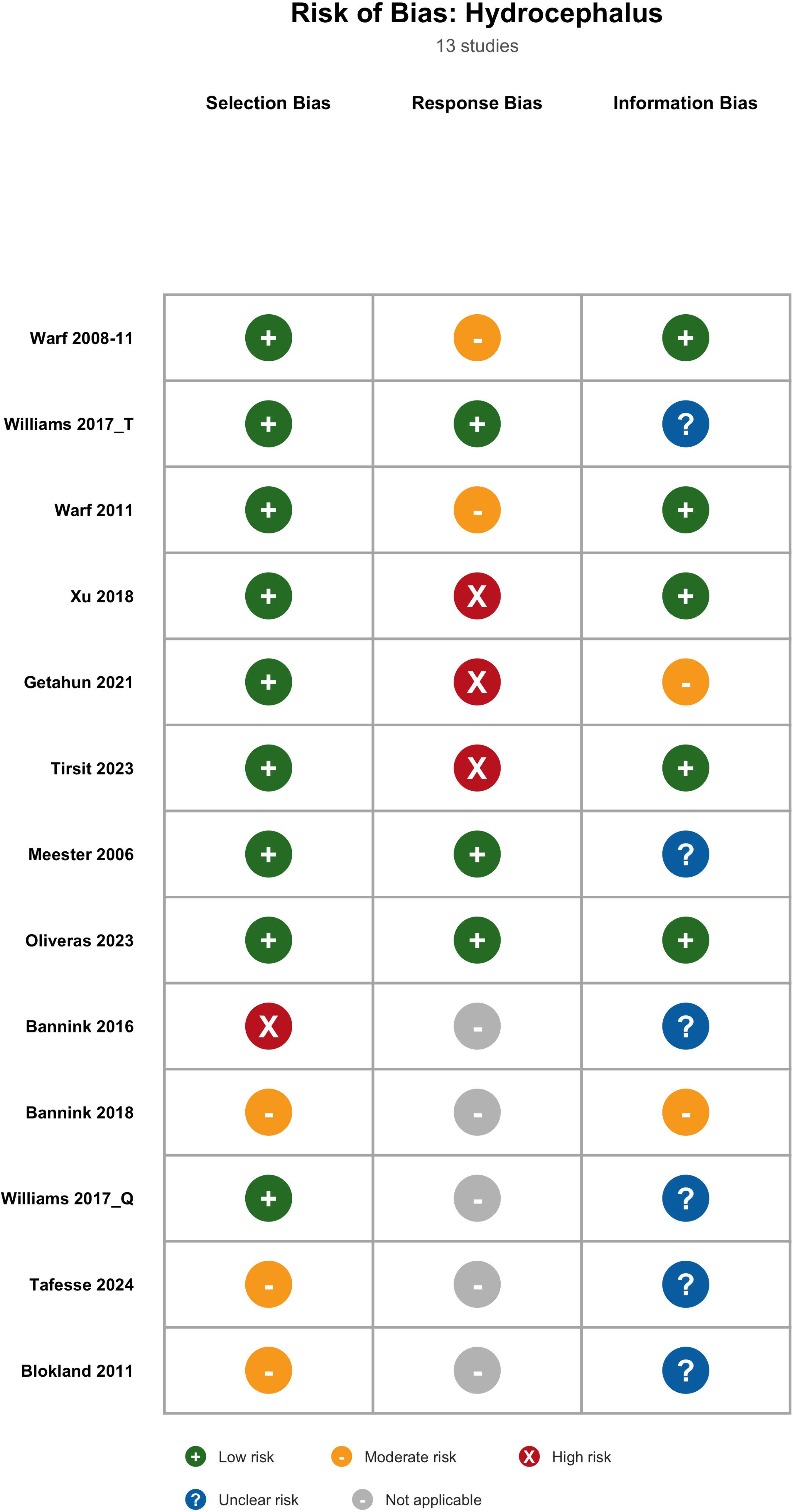

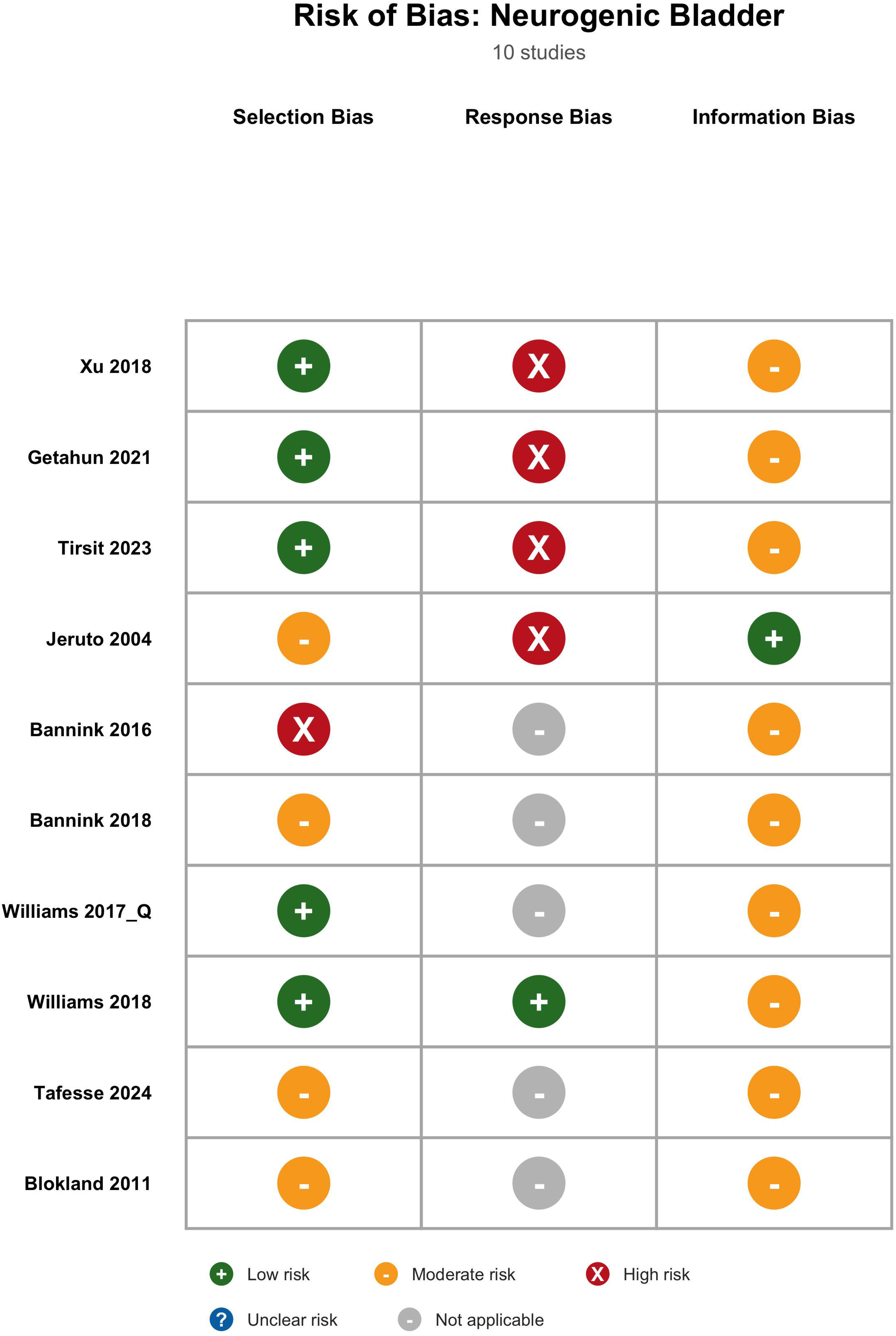

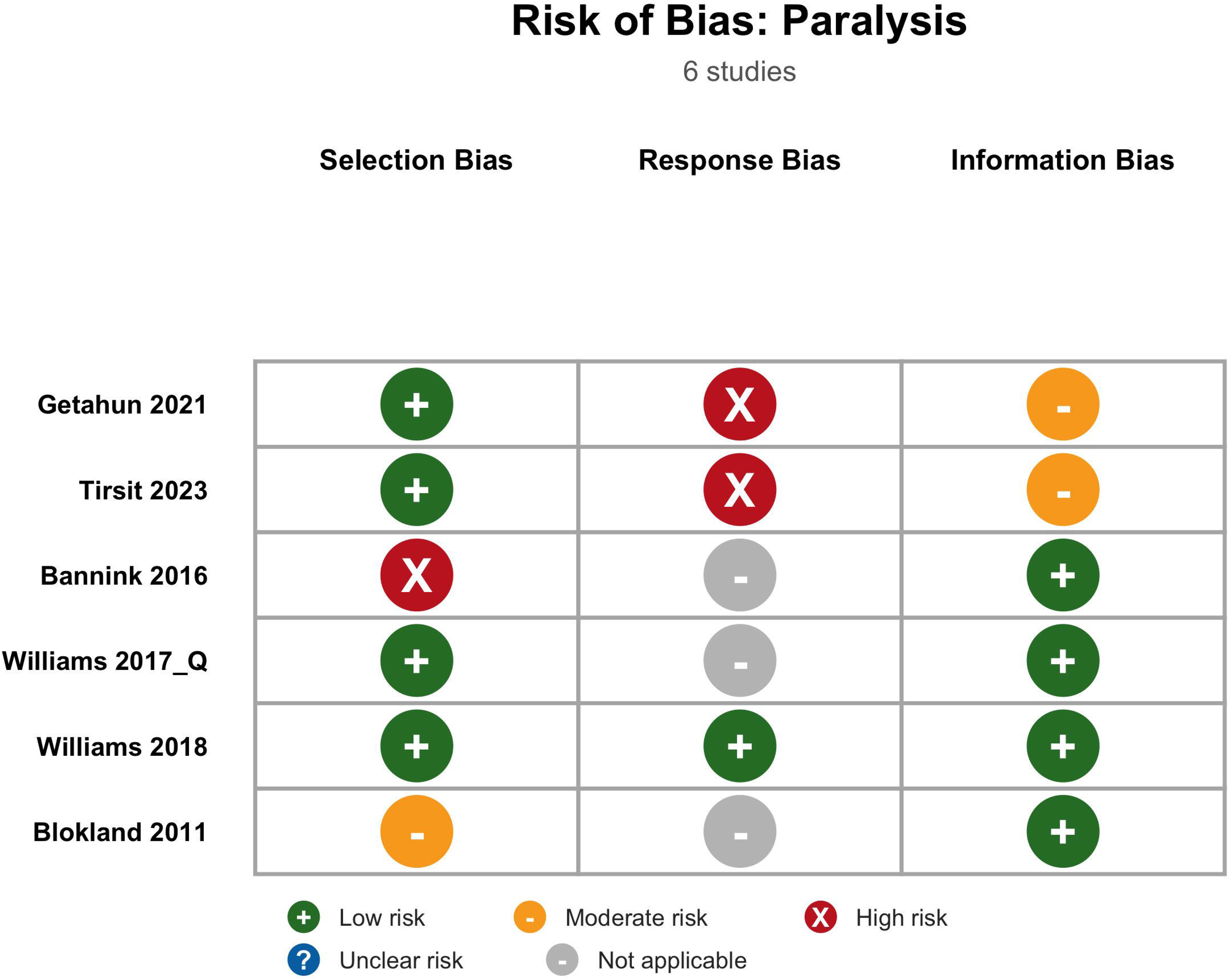

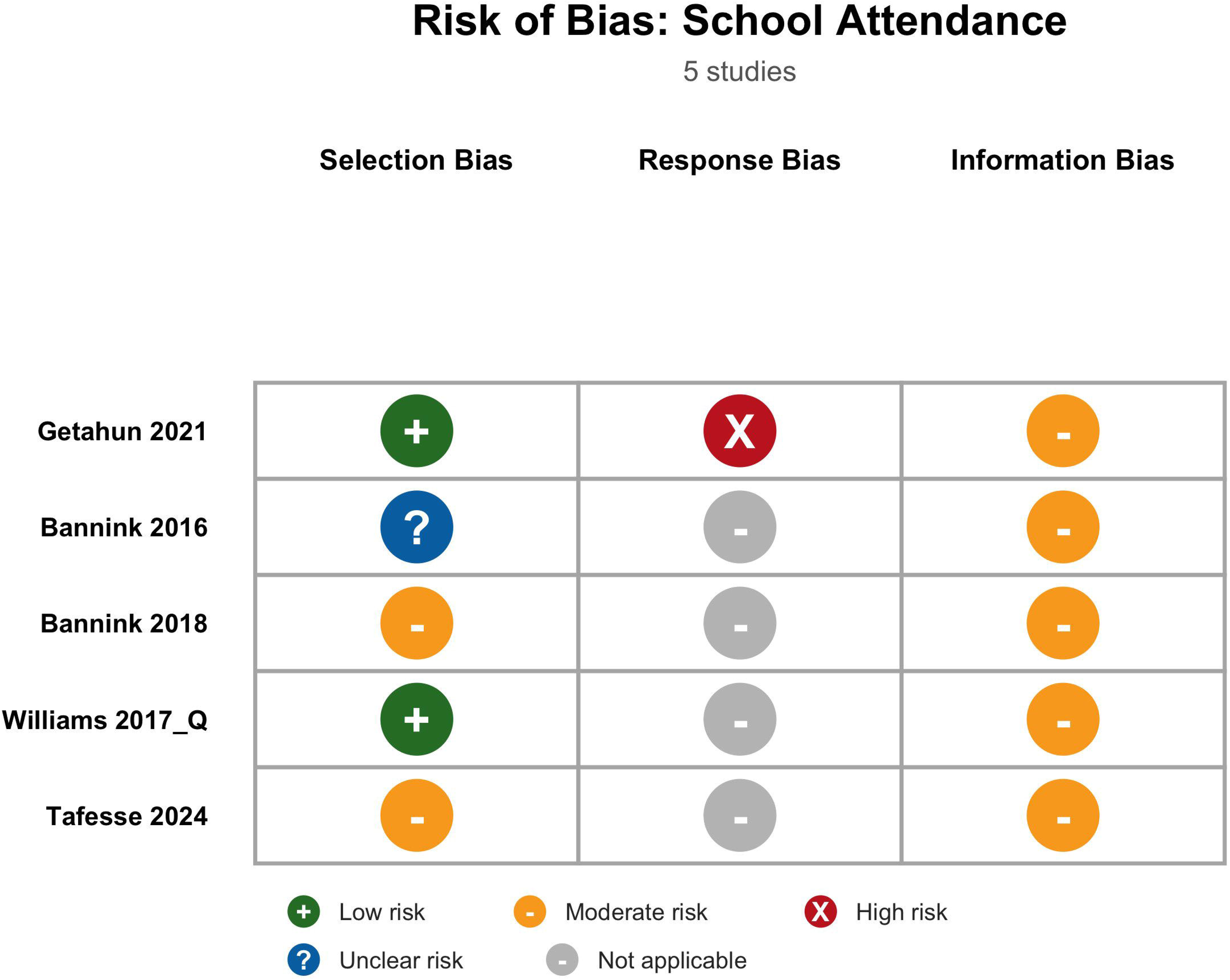

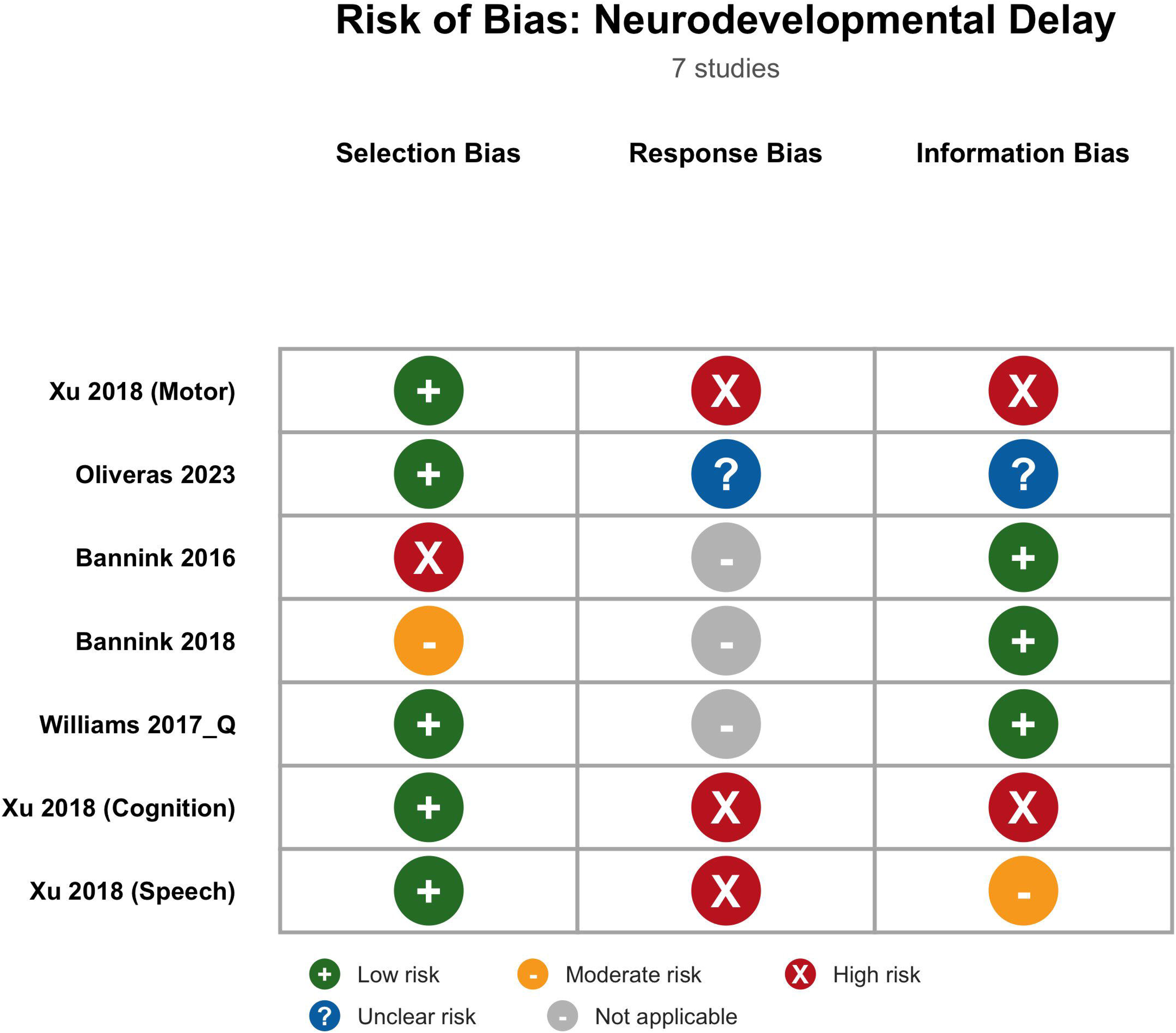

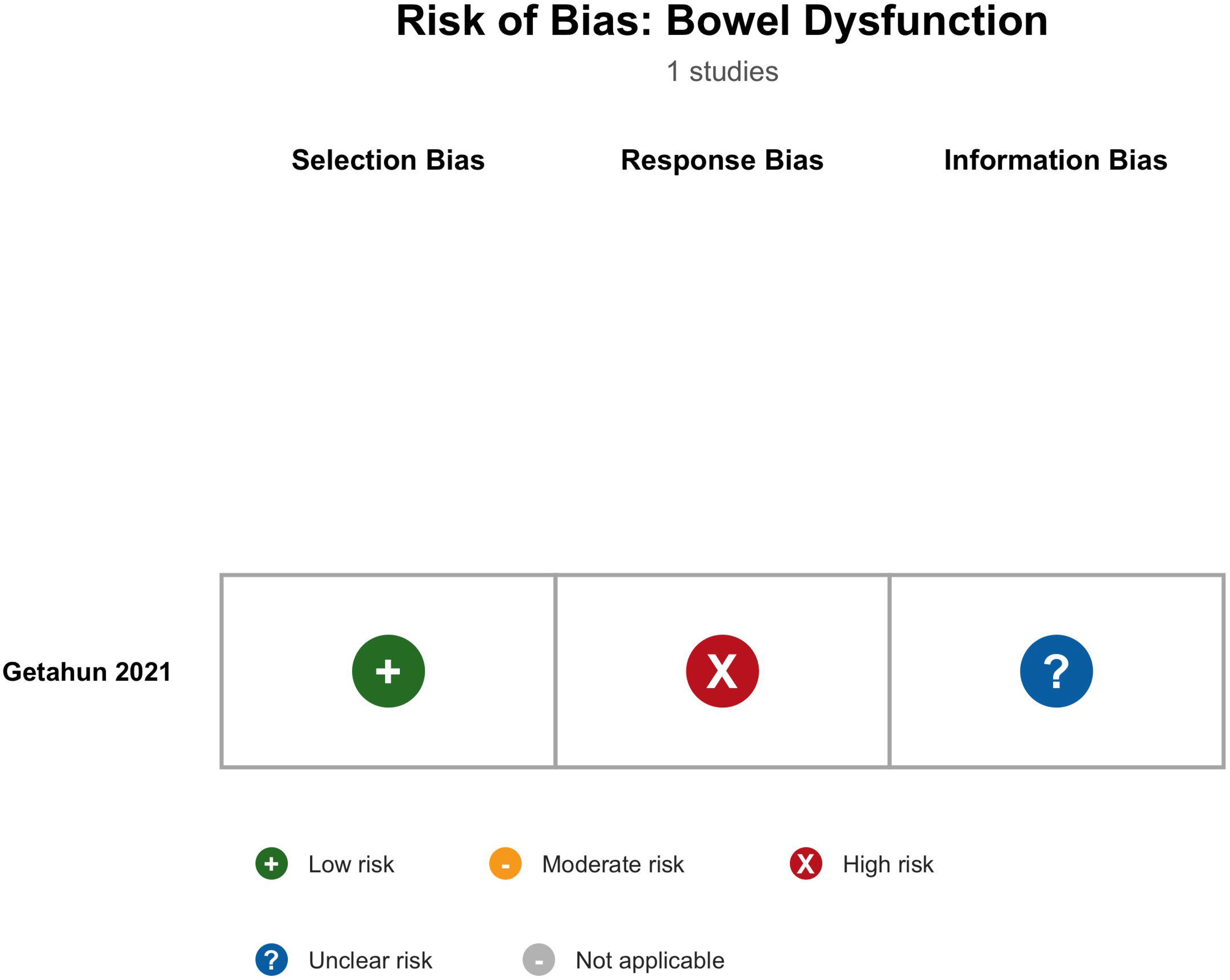

