## Supplementary Table 1 for "The Long-Term Outcomes of Neural Tube Defects in Eastern Africa: A Systematic Review and Meta-Analysis"

### 1. NTD Subtype Comparison: MMC vs Encephalocele

Date: 2025-11-21

#### 1.1. Statistical Test Results

##### 1.1.1. Mortality (13 months - 2 years):

Test: Chi-Square
Chi-Square (Q): 0.27
$I^2$ (Heterogeneity): 0 %
P-value: 0.6036
Significant: NO

##### 1.1.2. Hydrocephalus (all ages):

Test: Chi-Square
Chi-Square (Q): 0.36
$I^2$ (Heterogeneity): 0 %
P-value: 0.5508
Significant: NO

#### 1.2. Comparison Table

| Outcome | Myelomeningocele | Encephalocele | Test | Q | I_squared | P_value |
|---|---|---|---|---|---|---|
| Mortality (13m-2y) | 18% (57/317) | 21.1% (19/90) | Chi-Square | 0.27 | 0% | 0.6036 |
| Hydrocephalus | 38.5% (180/467) | 34.5% (30/87) | Chi-Square | 0.36 | 0% | 0.5508 |
