## Supplementary Text 2 for "The Long-Term Outcomes of Neural Tube Defects in Eastern Africa: A Systematic Review and Meta-Analysis"

**Systematic Review Protocol for Prospero**

#### Background

Neural tube defects (NTDs), also known as spinal dysraphism, are malformations that arise in the brain and spine of a developing foetus. They occur due to failure of the neural tube to close during the first stage of foetal development, usually up to four weeks from conception, and are induced by a complex interplay of genetic and environmental factors. NTDs are the most prevalent neurological anomalies in Africa [1]. Within sub-Saharan Africa (sSA), they are reported to be particularly common in the eastern African (EA) region [2], with an incidence of 3 in every 1000 births (equating to more than 1000 babies born with NTDs in the EA region each year).

NTDs are preventable through supplementation, or fortification of foods with folic acid [3-5]. Nevertheless, reducing the incidence of NTDs has been a challenge in low- and middle-income countries (LMICs). This is in part due to the high number of unplanned pregnancies in this region [6], coupled with insufficient pre-pregnancy folic acid supplementation. Furthermore, limited access to folic acid fortified foods in LMICs, as well as a lack of prenatal folic acid [7] due to delays in seeking antenatal care [8], further increases the burden of NTDs.

Given the scale of the problem, defining the long-term outcomes of children born with NTDs is an important step to informing the health care needs and continuing care among affected children. This need for improved healthcare and ongoing support was highlighted as an important research area by the World Health Assembly; in a recent forum [9], a call was made for newborn screening, diagnosis and management of birth anomalies with a focus on LMICs [10].

NTDs may be categorised into two groups. The first includes defects when the neural tube fails to close at the top. This affects brain development and leads to very severe but less common types of NTD, notably anencephaly, encephalocele and iniencephaly. Most children born with these types of NTD only survive for a few hours or are stillborn, except for those with encephalocele whose survival has become better over time [11]. Children with encephalocele may have an intelligent quotient (I.Q) above 90 [12], despite having special education needs, as well as physical disability and social development impairment. Furthermore, they may experience seizures, vision problems, a small head, facial and skull abnormalities, as well as uncoordinated movements.

The second group of NTD include the most prevalent NTDs [13], which occur when the neural tube fails to close completely and some of the bones of the spine do not close, resulting in spina bifida. Subgroups include spina bifida occulta, closed neural tube defect, meningocele and most commonly myelomeningocele [14]. Typically, children with spina bifida have few or no symptoms, however most children with meningocele or myelomeningocele experience varied outcomes including paralysis of the lower limbs, urinary and bowel dysfunction, hydrocephalus, and intellectual disabilities [15].

Previous systematic reviews have considered long-term survival of varied birth anomalies and showed a marked improvement in survival rates over the last two decades [16], based on mortality data in the under 5s. Nevertheless, a recent systematic estimate of the global, regional, and national under-5 mortality burden attributable to birth anomalies, inferred an underestimation of estimates in LMICs [10]; because of a limitation in the reporting of mortality due to birth defects, commonly obtained through verbal autopsy studies. Similarly, few studies are available with linked follow up of children beyond infancy, necessitating a thorough evaluation of the burden.

Therefore, we plan to undertake a systematic review and meta-analysis to investigate the long-term health outcomes associated with NTDs within the EA region, as an important and necessary step for developing healthcare policies in this field. To the best of our knowledge, this will be the first systematic review on long term outcomes of NTDs within the EA region.

**Review Question**

What are the long-term outcomes following NTDs among children in the eastern African region?

The objectives of this review are:

1. To analyse the survival rates of individuals with at least one NTD up to 18 years old.
2. To estimate the proportion of individuals with NTDs with long-term lower limb weakness or paralysis, bladder and bowel dysfunction, hydrocephalus, seizures, small head, facial and skull abnormalities and uncoordinated movements (exceeding 1 year of life up to 18 years).
3. To estimate the proportion of individuals with NTDs who attend school and those with special education needs (exceeding 3 years of life up to 18 years).
4. To describe the proportion of children with NTDs who have neurodevelopmental delays in vision, cognition, speech, hearing and motor function a (among individuals exceeding 1 year of life up to 18 years).

**Searches**

Publications of relevant studies will be identified using the following databases: PubMed/Medline, PubMed Central, Cochrane Library, Web of Science and Africa Index Medicus, Reference lists of identified articles and systematic reviews within the theme will also be scanned.

The language will be restricted in English.

**Search Strategy**

The search strategy is detailed in the appendix

**Condition or domain being studied**

The review will assess the long-term outcomes of NTDs including spina bifida, closed neural defect, meningocele, myelomeningocele, anencephaly, encephalocele and iniencephaly.

**Population**

*Inclusion*: Individuals under 18 years old born with a NTD.

*Exclusion:*

i. Individuals aged 18 years or older.
ii. Individuals under 18 years old without a NTD diagnosis at birth.

**Exposure**

*Inclusion*: Individuals under 18 years old who were diagnosed with any of the NTDs, or combination of a NTD with any anomalies at birth.

*Exclusion*: Individuals with other birth anomalies.

**Types of study to be included**

*Inclusion*: Studies with the below criteria, will be included.

1. Primary peer-reviewed study that reports one or a combination of the following outcomes.

i. Long-term (exceeding 1 year of life) survival of individuals born with NTDs
ii. Lower limb weakness or paralysis
iii. Uncoordinated movements
iv. Malfunction of the urine and bowel organs
v. Hydrocephalus
vi. Seizures
vii. Small head
viii. Facial and skull abnormalities
ix. Presence of special education needs
x. Neurodevelopmental delays (Vision, cognition, speech, hearing and motor skills)
xi. School attendance.

2. Birth cohorts of individuals born with a NTD or report on individuals with a NTD separately.
3. Case control, cohort and cross-sectional studies.
4. Involving humans only.

*Exclusion*: Studies not meeting the above standard and:

*1.* Involve only adults
*2.* Do not report separately on individuals with NTDs
*3.* Focus on other causes of the outcomes
*4.* Letters, books, editorials

**Context**
Study settings limited to the eastern Africa region

**Exposure of interest,**
Individuals born with a NTD which include:

- Spina bifida occulta
- Closed neural tube defect
- Meningocele
- Myelomeningocele
- Anencephaly
- Encephalocele
- Iniencephaly

**Measures of effect**

1. For studies with no control group
   Proportions or percentages (for dichotomous outcomes) and means or medians for numerical outcomes.
2. For studies with control group
   Hazard ratios (HR) for survival, odds ratios (OR) for other dichotomous outcomes, and standardised mean differences for numerical outcomes.

**Additional outcomes**
None

**Data extraction**

1. ***Selection of studies*** – titles/abstracts will be reviewed independently by two reviewers in this field, according to the specified inclusion/exclusion criteria and differences resolved through consensus discussion. Full text screening will be performed independently by two reviewers for inclusion and disagreements resolved through consensus discussion. If an agreement cannot be reached, a third reviewer will be consulted until a consensus is reached. When multiple reports of a study are identified these will be treated as a single study, but reference made to all publications. The Rayyan software system [17] will be used to facilitate the screening process.
2. ***Data Extraction*** – this will be performed by one reviewer; a second reviewer will independently check the data extraction forms for accuracy. Any disagreements will be noted and resolved by consensus or by arbitration from an additional independent researcher. Data will be extracted into a Microsoft excel. Pilot studies will be performed for the study selection and data extraction stages.

Six categories of information will be recorded:

1. Paper details
   a) Publication title

b) Author
c) Date
d) Journal Id

2. Population Characteristics

a) Country of study
b) Region type (rural/urban)
c) Number of cases with the condition –
    - Spina bifida occulta
    - Closed neural tube defect
    - Meningocele
    - Myelomeningocele
    - Anencephaly
    - Encephalocele
    - Iniencephaly
d) Age at baseline and follow up
e) Sex

3. Study design
    a) Case-control
    b) Cohort
    c) Cross sectional
4. Outcome(s)
    a) Mortality
    b) Lower limb weakness or paralysis
    c) Uncoordinated movements
    d) Urinary and bowel malfunction
    e) Hydrocephalus
    f) Seizures
    g) Small head
    h) Facial and skull abnormalities
    i) Special education needs
    j) Neurodevelopmental delays (Vision, speech, hearing, cognition and motor)
    k) School attendance
5. Results

a. Primary results; proportions, means, OR, incidence rate ratio (IRR), risk ratio (RR), risk difference (RD), HR.
b. Subgroup analysis
    i. Anencephaly, encephalocele, iniencephaly, spina bifida occulta, closed neural tube defect, meningocele and myelomeningocele.

ii. Age group analyses; toddlerhood, preschoolers, middle childhood/prepubertal and adolescence.

**Risk of bias assessment**

The risk of bias in the included studies will be assessed to address these specific domains.

1. Bias due to confounding
2. Bias arising from measurement of the exposure
3. Bias in selection of participants into the study
4. Bias due to post-exposure intervention
5. Bias due to missing data
6. Bias in the measurement of the outcome
7. Bias in selection of the reported result

Using the risk of bias in non-randomised studies - of exposure (ROBINS-E) tool, the review author will administer the tool to each included study, and record supporting information and justifications for judgements of risk of bias for each domain. Overall risk will be measured as (low risk; some concerns; high risk; very high risk).

**Strategy for Data Synthesis**

A clear descriptive summary of the primary studies and study characteristics will be done initially, and an indication of study quality will be recorded. For studies with no control group, we will present the percentages / means; for studies with a control group, we will present the HR (survival), OR (other dichotomous outcomes), or standardised mean differences (numerical outcomes). For studies that present other measures of effect (IRR, RR, RD), we will calculate an OR where possible using the published results. For studies showing sufficient homogeneity in terms of participants age, NTD subtype distribution and outcomes, we will conduct meta-analyses using random effects models with inverse-variance weighting to pool the estimates.

The extent of statistical heterogeneity will be examined using I-squared statistics, and this will be visualized on a forest plot. To assess the effect of a single study on the metaanalysis estimate, a leave-one-out sensitivity analysis will be done. Moreover, where there are sufficient studies funnel plots will be utilized to illustrate potential publication bias graphically. Where the number of studies or data is insufficient, or where studies are too heterogeneous for meta-analysis, a narrative analysis will be carried out. This will provide analysis of the outcomes within studies and a comprehensive evaluation of the validity of the evidence.

**Analysis of Subgroups**

Where the included number of studies which report results by subgroup is large enough, subgroup analysis will be performed for:

- Type of NTD (spina bifida, anencephaly, encephalocele, iniencephaly, meningocele, myelomeningocele, closed neural tube defect).
- Age group analyses; toddlerhood, preschoolers, middle childhood/prepubertal and adolescence.

### Appendix

*Appendix A*

**Table 1.** Criteria for including or excluding papers from this systematic review.

| Include | Exclude |
|---|---|
| 1. Full-text papers published in a peer-reviewed journal. | 1. Title, abstract or conference proceedings only or published in a non-peer reviewed journal (book, newspaper, letter or website). |
| 2. Primary data using observational study designs, (looking at outcomes related to NTDs.) These may include cohort, cross-sectional studies and case-control studies. | 2. All other study types including individual case studies, case series, expert opinion, qualitative studies, letters and editorials and secondary data from reviews. |
| 3. Exposure: NTDs | 3. Other birth anomalies |
| 4. Outcome measured: mortality, lower limb weakness or paralysis, uncoordinated movements, urinary and bowel malfunction, hydrocephalus, seizures, small head, facial and skull abnormalities, special education needs and neurodevelopmental delays (vision, speech, hearing, cognition and motor). | 4. Other outcomes not associated with NTDs. |
| 5. Study of humans. | 5. Study of animals or cell studies. |

*Appendix B*

**The search strategy**

The search will be for studies evaluating longterm effects of NTDs (Spina bifida occulta, closed neural tube defect, meningocele, myelomeningocele, anencephaly, encephalocele and iniencephaly). Published from beginning to present in PubMed/Medline, PubMed Central, Cochrane Library, Web of Science and Africa Index Medicus.

**Pubmed**

("Neural tube defects" [MeSH] OR "Spinal dysraphisms" [MeSH] "Spina bifida occulta" [ MeSH] OR "Meningocele" [MeSH] OR "Meningomyelocele" [MeSH] OR "Anencephaly" [M eSH] OR "Encephalocele" [MeSH] OR "Neural tube defects" [Title/abstract] OR "Spinal d ysraphisms" [Title/abstract] OR "NTDs" [Title/abstract]" OR "Spina bifida occulta" [Title /abstract] OR "Closed neural tube defect" [Title/abstract] OR "Meningocele" [Title/abstr act] OR "Myelomeningocele" [Title/abstract] OR "Anencephaly" [Title/abstract] OR "Enc ephalocele" [Title/abstract] OR "Iniencephaly" [Title/abstract])

AND

("Long term adverse effects" [MeSH] OR "Persistent vegetative state" [MeSH] OR "Long t erm adverse effects" [Title/abstract] OR "Persistent vegetative state" [Title/abstract] O R "Long term effects" [Title/abstract] OR "Long term impacts" [Title/abstract] OR "Long term outcome" [Title/abstract] OR "Long term sequelae" [Title/abstract] "Long*standin g effects" [Title/abstract] OR "Long standing impacts" [Title/abstract ] OR "Long standin g outcome" [Title/abstract] OR "Long*standing sequelae" [Title/abstract] OR "Permane nt effects" [Title/abstract] OR "Permanent impacts" [Title/abstract] OR "Permanent out come" [Title/abstract] OR "Permanent sequelae" [Title/abstract] OR "Extended effects" [Title/abstract] OR "Extended impacts" [Title/abstract] OR "Extended outcome" [Title/a bstract] OR "Extended sequelae" [Title/abstract])

OR

("Survival" [MeSH] OR "Mortality" [MeSH] OR "Child Mortality" [Mesh] OR "Infant Mortality" [MeSH] OR "Paralysis" [MeSH] OR "Urinary Incontinence" [MeSH] OR "Urinar y bladder neurogenic" [MeSH] OR "Constipation" [MeSH] OR "Hydrocephalus" [MeSH] O R "Intellectual disability" [MeSH] OR "Persons with disabilities" [MeSH] OR "Children w ith disabilities" [MeSH] OR "Cognitive dysfunction" [MeSh] OR "Ataxia" [MeSH] OR "Micr ocephaly" [MeSH] OR "Seizures" [MeSH] OR "Surviv*" [Title/abstract] OR "Mortality" [Ti tle/abstract] OR "Child Mortality" [Title/abstract] OR "Infant Mortality" [Title/abstract] OR "Paralysis" [Title/abstract] OR "Lower limb weakness" [Title/abstract] OR "Bladder dysfunction" [Title/abstract] OR "Urin* Incontinence" [Title/abstract]) OR "Urinary blad der neurogenic" [Title/abstract] OR "Bowel dysfunction" [Title/abstract] OR "Constipati on" [Title/abstract] OR "Faecal incontinence" [Title/abstract] OR "Hydrocephalus" [Title /abstract] OR "Special education needs" [Title/abstract] OR "Disab*" [Title/abstract] OR "Intellectual disability" [Title/abstract] OR "Persons with disabilities" [Title/abstract] O R "Children with disabilities" [Title/abstract] OR "Impairment" [Title/abstract] OR "Ha ndicapped" [Title/abstract] OR "Cognitive dysfunction" [Title/abstract] OR "Ataxia" [Titl e/abstract] OR "Uncoordinated movements" [Title/abstract] OR "Seizures" [Title/abstra ct] OR "Microcephaly" [Title/abstract] OR "Small head" [Title/abstract] OR "Facial abno rmalities" [Title/abstratct] and "Skull abnormalities" [Title/abstract] OR "Neurodevelop mental delays" [Title/abstract] OR "School attendance" [Title/abstract])

AND

("Africa, Eastern" [MeSH] OR "Africa south of the sahara" [MeSH] OR "Africa" [MeSH] OR "Rwanda" [MeSH] OR "Mauritius" [MeSH] OR "Mozambique" [MeSH] OR "Zimbabwe" [ MeSH] OR "Malawi" [MeSH] OR "Zambia" [MeSH] OR "East*Africa" [Title/abstract] OR " Africa eastern" [Title/abstract] OR "Sub saharan Africa" [Title/abstract] OR "Africa" [ Tit le/abstract] OR "Rwanda" [Title/abstract] OR "Mauritius" [ Title/abstract] OR "Mozamb ique" [Title/abstract] OR "Zimbabwe" [Title/abstract] OR "Malawi" [Title/abstract] OR " Zambia" [Title/abstract] OR "Mayotte " [Title/abstract] OR "Kenya" [Title/abstract] OR " Tanzania" [Title/abstract] OR "Uganda" [Title/abstract] OR "Burundi" [Title/abstract] O R "Ethiopia" [Title/abstract] OR "South Sudan" [Title/abstract] OR "Somali" [Title/abstr act] OR "Eriteria" [Title/abstract] OR "Seychelles" [Title/abstract] OR "Sudan" [Title/abs tract] OR "Comoros" [Title/abstract] OR "Djibout" [Title/abstract] OR "Madagasca" [Title/abstract])

**Medline**
("Neural tube defects" [MeSH] OR "Spinal dysraphisms" [MeSH] "Spina bifida occulta" [ MeSH] OR "Meningocele" [MeSH] OR "Meningomyelocele" [MeSH] OR "Anencephaly" [M eSH] OR "Encephalocele" [MeSH] OR "Neural tube defects" [Keyword] OR "Spinal dysrap hisms" [ Keyword] OR "NTD*" [ Keyword]" OR "Spina bifida occulta" [ Keyword] OR "Clo sed neural tube defect" [Keyword] OR "Meningocele" [ Keyword] OR "Meningomyelocel e" [ Keyword] OR "Anencephaly" [Keyword] OR "Encephalocele" [ Keyword] OR "Inience phaly" [ Keyword])

AND
("Long term adverse effects" [MeSH] OR "Persistent vegetative state" [MeSH] OR "long term adverse effects" [Keyword] OR "Persistent vegetative state" [Keyword] OR "Long t erm effects" [Keyword] OR "Long term impacts" [ Keyword ] OR "Long term Outcome" [ Keyword] OR "Long term sequelae" [Keyword] "Long*standing effects" [Keyword] OR "L ong*standing impacts" [Keyword] OR "Long standing outcome" [Keyword ]OR "Long*st anding sequelae" [Keyword] OR "Permanent effects" [Keyword] OR "Permanent impacts " [Keyword] OR "Permanent outcome" [Keyword] OR "Permanent sequelae" [Keyword] OR "Extended effects" [Keyword] OR "Extended impacts" [Keyword] OR "Extended outc ome" [Keyword] OR "Extended sequelae" [Keyword])

OR
("Survival" [MeSH] OR "Mortality" [MeSH] OR "Child Mortality" [Mesh] OR "Infant Morta lity" [MeSH] "Paralysis" [MeSH] OR "Urinary Incontinence" [MeSH]) OR "Urinary bladde r neurogenic" [MeSH] OR "Constipation" [MeSH] OR "Faecal incontinence" [MeSH] OR " Hydrocephalus" [MeSH] OR "Intellectual disability" [MeSH] OR "Disabled persons" [MeS H] OR "Disabled Children" [MeSH] OR "Cognitive dysfunction" [MeSh] OR "Ataxia" [MeS H] OR "Seizures" [MeSH] OR "Microcephaly" [MeSH] OR "Survival" [Keyword] OR "Mort ality" [Keyword] OR "Child Mortality" [Keyword] OR "Infant Mortality" [Keyword] OR "P aralysis" [Keyword] OR "Lower limb weakness" [ Keyword] OR "Bladder dysfunction" [K eyword] OR "Urine Incontinence" [Keyword] OR "Urinary bladder neurogenic" [Keywor d] OR "Bowel dysfunction" [Keyword] OR "Constipation" [ Keyword] OR "Faecal inconti nence" [ Keyword] OR "Hydrocephalus" [ Keyword] OR "Special education needs" [Keyw ord] OR "Intellectual disability" [Keyword] OR "Disabled persons" [Keyword] OR "Disabl ed Children" [Keyword] OR "Impairment" [ Keyword] OR "Handicapped" [Keyword] OR "Cognitive dysfunction" [Keyword] OR "Ataxia" [Keyword] OR "Uncoordinated moveme nts" [Keyword] OR "Seizures" [Keyword] OR "Microcephaly" [Keyword] OR "Small head"

[Keyword] OR "Facial abnormalities" [Keyword] and "Skull abnormalities" [Keyword] OR "Neurodevelopmental delays" [Keyword] OR "School attendance" [Keyword] )

AND
("Africa Eastern" [MeSH] OR "Africa south of the Sahara" [MeSH] OR "Africa" [MeSH] OR "Rwanda" [MeSH] OR "Mauritius" [MeSH] OR "Mozambique" [MeSH] OR "Zimbabwe" [MeSH] OR "Malawi" [MeSH] OR "Zambia" [MeSH] OR "Africa Eastern " [Keyword] OR "Africa south of the Sahara" [Keyword] OR "Africa" [Keyword] OR "Rwanda" [Keyword] OR " Mauritius" [Keyword] OR "Mozambique" [Keyword] OR "Zimbabwe" [Keyword] OR "Malawi" [Keyword] OR "Zambia" [ Keyword] OR "Mayotte" [Keyword] OR "Kenya" [ Keyword] OR "Tanzania" [Keyword] OR "Uganda" [Keyword] OR "Burundi" [Keyword] OR "Ethiopia" [Keyword] OR "South Sudan" [Keyword] OR "Somali" [Keyword] OR "Eriteria" [Keyword] OR "Seychelles" [Keyword] OR "Sudan" [Keyword] OR "Comoros" [Keyword] OR "Djibout" [Keyword] OR "Madagasca" [Keyword])

**Cochrane Library**
#1= Neural tube defects [MeSH] OR Spinal dysraphisms [MeSH] Spina bifida occulta [MeSH] OR Meningocele [MeSH] OR Myelomeningocele [MeSH] OR Anencephaly [MeSH] OR Encephalocele [MeSH]

#2= "Neural tube defects" [Title/abstract/keyword] OR "NTD" [Title/abstract/keyword] OR "Spinal dysraphisms" [Title/abstract/keyword] OR "Spina bifida occulta" [Title/abstract/keyword] OR "Closed neural tube defect" [Title/abstract/keyword] OR "Meningocele" [Title/abstract/keyword] OR "Myelomeningocele" [Title/abstract/keyword] OR "Anencephaly" [Title/abstract/keyword] OR "Encephalocele" [Title/abstract/keyword] OR "Iniencephaly" [Title/abstract/keyword]

#3= #1 AND #2

#4= "Long term adverse effects" [Title/abstract/keyword] OR "Persistent vegetative state" [Title/abstract/keyword] OR "Long term effects" [Title/abstract/Keyword] OR "Long term impacts" [Title/abstract/keyword] OR "Long term outcome" [Title/abstract/keyword] OR "Long term sequelae" [Title/abstract/keyword] OR "Long standing effects" [Title/abstract/keyword] OR "Long standing impact" [Title/abstract/keyword] OR "Long standing outcome" [Title/abstract/keyword] OR "Long standing sequelae" [Title/abstract/keyword] OR "Permanent effects" [Title/abstract/keyword] OR "Permanent impact" [ Title/abstract/keyword] OR "Permanent outcome" [Title/abstract/keyword] OR "Permanent sequelae" [Title/abstract/keyword] OR "Extended effects" [Title/abstract/keyword] OR "Extended impact" [Title/abstract/keyword] OR "Extended outcome" [Title/abstract/keyword] OR "Extended sequelae" [Title/abstract/keyword]

#5 = #3 AND #4

#6 Survival [MeSH] OR Mortality [MeSH] OR "Survival" [Title/abstract/keyword] OR "Mortality" [Title/abstract/keyword] OR "Child Mortality" [Title/abstract/keyword] OR "Infant Mortality" [Title/abstract/keyword] OR "Paralysis" [Title/abstract/keyword] OR "Lower limb weakness" [Title/abstract/keyword] OR "Bladder dysfunction" [Title/abstract/keyword] OR "Urine Incontinence" [Title/abstract/keyword] OR "Urinary bladder neurogenic" [Title/abstract/keyword] OR "Bowel dysfunction" [Title/abstract/keyword]

OR "Constipation" [Title/abstract/keyword] OR "Faecal incontinence" [Title/abstract/keyword] OR "Hydrocephalus" [Title/abstract/keyword] OR "Special education needs" [Title/abstract/keyword] OR "Disability" [Title/abstract/keyword] OR "Intellectual disability" [Title/abstract/keyword] OR "Disabled persons" [Title/abstract/keyword] OR "Disabled Children" [Title/abstract/keyword] OR "Impairment" [Title/abstract/keyword] OR "Handicapped" [Title/abstract/keyword] OR "Cognitive dysfunction" [Title/abstract/keyword] OR "Ataxia" [Title/abstract/keyword] OR "Uncoordinated movements" [Title/abstract/keyword] OR "Seizure" [Title/abstract/keyword] OR "Microcephaly" [Title/abstract/keyword] OR "Small head" [Title/abstract/keyword] OR "Facial abnormalities" [Title/abstract/keyword] and "Skull abnormalities" [Title/abstract/keyword] OR "Neurodevelopmental delays" [Title/abstract/keyword] OR "School attendance" [Title/abstract/keyword]

#7 = #3 AND #6

#8= #5 OR #7

#9= Africa, Eastern [MeSH] OR Africa South of the Sahara [MeSH] OR Africa [MeSh] OR "Africa, Eastern" [Title/abstract/keyword] OR "Africa South of the Sahara" [Title/abstract/keyword] OR "Africa" [Title/abstract/keyword] OR "Kenya" [Title/abstract/keyword] OR "Tanzania" [Title/abstract/keyword] OR "Uganda" [Title/abstract/keyword] OR "Burundi" [Title/abstract/keyword] OR "Ethiopia" [Title/abstract/keyword] OR "South Sudan" [Title/abstract/keyword] OR "Somali" [Title/abstract/keyword] OR "Rwanda" [Title/abstract/keyword] OR "Eriteria" [Title/abstract/keyword] OR "Mauritius" [Title/abstract/keyword] OR "Seychelles" [Title/abstract/keyword] OR "Sudan" [Title/abstract/keyword] OR "Comoros" [Title/abstract/keyword] OR "Djibout" [Title/abstract/keyword] OR "Mozambique" [Title/abstract/keyword] OR "Zimbabwe" [Title/abstract/keyword] OR "Malawi" [Title/abstract/keyword] OR "Zambia" [Title/abstract/keyword] OR "Madagasca" [Title/abstract/keyword] OR "Mayotte" [Title/abstract/keyword]

#10=#8 AND #9

**Web of Science**

1#=Topic (Ts) "Neural tube defects" OR "NTDs" OR "Spinal dysraphisms" OR "Spina bifida occulta" OR "Closed neural tube defect" OR "Meningocele" OR "Myelomeningocele" OR "Anencephaly" OR "Encephalocele" OR "Iniencephaly"

2#=Ts "Long term adverse effects" OR "Persistent vegetative state" OR "Long term effects" OR "Long term impacts" OR "Long term Outcome" OR "Long term sequelae" OR "Long standing effects" OR "Long standing impacts" OR "Long standing outcome" OR "Long standing sequelae" OR "Permanent effects" OR "Permanent impacts" OR "Permanent outcome" OR "Permanent sequelae" OR "Extended effects" OR "Extended impacts" OR "Extended outcome" OR "Extended sequelae"

3#=1# AND 2#

4#=Ts “Survival” OR “Mortality” OR "Child Mortality” OR "Infant Mortality" OR “Paralysis” OR “Lower limb weakness” OR “Bladder dysfunction” OR “Urinary Incontinence” OR “ Urinary bladder neurogenic” OR “Bowel dysfunction” OR “Constipation” OR “Faecal incontinence” OR “Hydrocephalus” OR “Special education needs” OR “Disability” OR “Intellectual disability” OR “Disabled persons” OR “Disabled Children” OR “Impairment” OR “Handicapped” OR “Cognitive dysfunction” OR “Ataxia” OR “Uncoordinated movements” OR “ Seizure” OR “Small head” OR “Microcephaly” OR “Facial abnormalities” OR “Skull abnormalities” OR “Neurodevelopmental delays” OR “School attendance”

#5=#1 AND #4

#6= #3 OR #5

7#= Ts “Africa Eastern” OR “East Africa” OR “Africa south of the Sahara” OR "Sub Sahara Africa" OR “Africa” OR “Kenya” OR “Tanzania” OR “Uganda” OR “Burundi” OR “Ethiopia” OR “South Sudan” OR “Somali” OR “Rwanda” OR “Eriteria” OR “Mauritius” OR “Seychelles” OR “Sudan” OR “Comoros” OR “Djibout” OR “Mozambique” OR “Zimbabwe” OR “Malawi” OR “Zambia” OR “Madagasca” OR “Mayotte”

#8= #6 AND #7

**African Index Medicus**
“Neural tube defects” [Title/abstract/subject] OR “NTDs” [Title/abstract/subject] OR “Spinal dysraphisms” [Title/abstract/subject] OR “Spina bifida occulta” [Title/abstract/subject] OR “Closed neural tube defect” [Title/abstract/subject] OR “Meningocele” [Title/abstract/subject] OR “Myelomeningocele” [Title/abstract/subject] OR “Anencephaly” [Title/abstract/subject] OR “Encephalocele” [Title/abstract/subject] OR “Iniencephaly” [Title/abstract/ subject]

AND
“Longterm adverse effects” [Title/abstract/subject] OR “Persistent vegetative state” [Title/abstract/subject] OR “Longterm impacts” [Title/abstract/subject] OR “Longterm outcome” [Title/abstract/Keyword] OR “Longterm sequelae” [Title/abstract/subject] OR “longstanding effects” [Title/abstract/subject] OR “Longstanding impacts” [Title/abstract/subject] OR “Longstanding outcome” [Title/abstract/subject] OR “Longstanding sequelae” [Title/abstract/subject] OR “Permanent effects” [Title/abstract/subject] OR “Permanent impacts” [Title/abstract/subject] OR “Permanent outcome” [Title/abstract/subject] OR “Permanent sequelae” [Title/abstract/subject] OR “Extended effects” [Title/abstract/subject] OR “Extended impacts” [Title/abstract/subject] OR “Extended outcome” [Title/abstract/ subject] OR “Extended sequelae” [Title/abstract/ subject]

OR
“Surviv*” [Title/abstract/subject] OR “Mortality” [Title/abstract/subject] OR "Child Mortality" [Title/abstract/subject] OR "Infant Mortality" [Title/abstract/subject] OR “Paralysis” [Title/abstract/subject] OR “Lower limb weakness” [Title/abstract/subject] OR “Bladder dysfunction” [Title/abstract/subject] OR “Urine Incontinence” [Title/abstract/subject] OR “Urinary bladder neurogenic” [Title/abstract/subject] OR “Bowel dysfunction” [Title/abstract/subject] OR “Constipation” [Title/abstract/subject] OR “Faecal incontinence” [Title/abstract/subject] OR “Hydrocephalus” [Title/abstract/subject] OR “Spec

ial education needs" [Title/abstract/ subject] OR "Disab*" [Title/abstract/subject] OR "I ntellectual disability" [Title/abstract/subject] OR "Disabled persons" [Title/abstract/ subject] OR "Disabled Children" [Title/abstract/subject] OR "Impairment" [Title/abstra ct/subject] OR "Handicapped" [Title/abstract/subject] OR "Cognitive dysfunction" [Title /abstract/subject] OR "Ataxia" [Title/abstract/subject] OR "Uncoordinated movements" [Title/abstract/subject] OR "Seizure" [Title/abstract/subject] OR "Microcephaly" [Title /abstract/subject] OR "Small head" [Title/abstract/subject] OR "Facial abnormalities" [ Title/abstract/subject] OR "Skull abnormalities" [Title/abstract/subject] OR "Neurodev elopmental delays" [Title/abstract/subject] OR "School attendance" [Title/abstract/sub ject]

AND
"Africa Eastern" [Title/abstract/subject] OR "Africa south of the sahara" [Title/abstract/ subject] OR "Africa" [Title/abstract/subject] OR "Kenya" [Title/abstract/subject] OR "Ta nzania" [Title/abstract/subject] OR "Uganda" [Title/abstract/subject] OR "Burundi" [Tit le/abstract/subject] OR "Ethiopia" [Title/abstract/subject] OR "South Sudan" [Title/abs tract/subject] OR "Somali" [Title/abstract/subject] OR "Rwanda" [Title/abstract/subjec t] OR "Eriteria" [Title/abstract/subject] OR "Mauritius" [Title/abstract/subject] OR "Sey chelles" [Title/abstract/subject] OR "Sudan" [Title/abstract/subject] OR "Comoros" [Tit le/abstract/subject] OR "Djibout" [Title/abstract/subject] OR 'Mozambique" [Title/abst ract/subject] OR "Zimbabwe" [Title/abstract/subject] OR "Malawi" [Title/abstract/subj ect] OR "Zambia" [Title/abstract/subject] OR "Madagasca" [Title/abstract/ subject] OR "Mayotte" [Title/abstract/ subject]
