## Supplementary Text 3 for "The Long-Term Outcomes of Neural Tube Defects in Eastern Africa: A Systematic Review and Meta-Analysis"

AND

(“Africa, Eastern” [MeSH] OR “Africa south of the sahara ” [MeSH] OR “Africa” [MeSH] O
R “Rwanda” [MeSH] OR “Mauritius” [MeSH] OR “Mozambique” [MeSH] OR “Zimbabwe” [
MeSH] OR “Malawi” [MeSH] OR “Zambia” [MeSH] OR “East*Africa” [Title/abstract] OR “
Africa eastern ” [Title/abstract] OR “Sub saharan Africa” [Title/abstract] OR “Africa” [ Ti
tle/abstract] OR “Rwanda ” [Title/abstract] OR “Mauritius” [ Title/abstract] OR “Mozam
bique” [Title/abstract] OR “Zimbabwe” [Title/abstract] OR “Malawi” [Title/abstract] OR
“Zambia” [Title/abstract] OR “Mayotte ” [Title/abstract] OR “Kenya” [Title/abstract] OR
“Tanzania” [Title/abstract] OR “Uganda” [Title/abstract] OR “Burundi” [Title/abstract]
OR “Ethiopia” [Title/abstract] OR “South Sudan” [Title/abstract] OR “Somali” [Title/abst
ract] OR “Eriteria” [Title/abstract] OR “Seychelles” [Title/abstract] OR “Sudan” [Title/ab
stract] OR “Commoros” [Title/abstract] OR “Djibout” [Title/abstract] OR “Madagasca”
[Title/abstract])

**Medline**
(“Neural tube defects” [MeSH] OR “Spinal dysraphisms” [MeSH] “Spina bifida occulta” [
MeSH] OR “Meningocele” [MeSH] OR “Meningomyelocele” [MeSH] OR “Anencephaly” [M
eSH] OR “Encephalocele” [MeSH] OR “Neural tube defects” [Keyword] OR “Spinal dysrap
hisms” [ Keyword] OR “NTD*” [ Keyword]” OR “Spina bifida occulta” [ Keyword] OR “Clo
sed neural tube defect” [Keyword] OR “Meningocele” [ Keyword] OR “Meningomyelocel
e” [ Keyword] OR “Anencephaly” [Keyword] OR “Encephalocele” [ Keyword] OR “Inience
phaly” [ Keyword])

AND
(“Long term adverse effects” [MeSH] OR “Persistent vegetative state” [MeSH] OR “long
term adverse effects” [Keyword] OR “Persistent vegetative state” [Keyword] OR “Long t
erm effects” [Keyword] OR “Long term impacts” [ Keyword ]OR “Long term Outcome” [
Keyword] OR “Long term sequelae” [Keyword] “Long*standing effects” [Keyword] OR “L
ong*standing impacts” [Keyword] OR “Long standing outcome” [Keyword ]OR “Long*st
anding sequelae” [Keyword] OR “Permanent effects” [Keyword] OR “Permanent impacts
” [Keyword] OR “Permanent outcome” [Keyword] OR “Permanent sequelae” [Keyword]
OR “Extended effects” [Keyword] OR “Extended impacts” [Keyword] OR “Extended outc
ome” [Keyword] OR “Extended sequelae” [Keyword])

OR
(“Survival” [MeSH] OR “Mortality” [MeSH] OR "Child Mortality" [Mesh] OR "Infant Morta
lity" [MeSH] “Paralysis” [MeSH] OR “Urinary Incontinence” [MeSH]) OR “Urinary bladde
r neurogenic” [MeSH] OR “Constipation” [MeSH] OR “Faecal incontinence” [MeSH] OR “
Hydrocephalus” [MeSH] OR “Special education needs” [Title/abstract] “Intellectual disa
bility” [MeSH] OR “Disabled persons” [MeSH] OR “Disabled Children” [MeSH] OR “Cognit
ive dysfunction” [MeSh] OR “Ataxia” [MeSH] OR “Seizures” [MeSH] OR “Microcephaly” [
MeSH] OR “Survival” [Keyword] OR “Mortality” [Keyword] OR "Child Mortality" [Keywo
rd] OR "Infant Mortality" [Keyword] OR “Paralysis” [Keyword] OR “Lower limb weaknes
s” [ Keyword] OR “Bladder dysfunction” [Keyword] OR “Urine Incontinence” [Keyword]
OR “Urinary bladder neurogenic” [Keyword] OR “Bowel dysfunction” [Keyword] OR “Co
nstipation” [ Keyword] OR “Faecal incontinence” [ Keyword] OR “Hydrocephalus” [ Key
word] OR “Special education needs” [Keyword] OR “Intellectual disability” [Keyword] O
R “Disabled persons” [Keyword] OR “Disabled Children” [Keyword] OR “Impairment” [
Keyword] OR “Handicapped” [Keyword] OR “Cognitive dysfunction” [Keyword] OR “Ata
xia” [Keyword] OR “Uncoordinated movements” [Keyword] OR “Seizures” [Keyword] O

#7 = #3 AND #6

#8= #5 OR #7

#9= Africa, Eastern [MeSH] OR Africa South of the Sahara [MeSH] OR Africa [MeSh] OR “ Africa, Eastern” [Title/abstract/keyword] OR “Africa South of the Sahara” [Title/abstrac t/keyword] OR “Africa” [Title/abstract/keyword] OR “Kenya” [Title/abstract/keyword] OR “Tanzania” [Title/abstract/keyword] OR “Uganda” [Title/abstract/keyword] OR “B urundi” [Title/abstract/keyword] OR “Ethiopia” [Title/abstract/keyword] OR “South Su dan” [Title/abstract/keyword] OR “Somali” [Title/abstract/keyword] OR “Rwanda” [Tit le/abstract/keyword] OR “Eriteria” [Title/abstract/keyword] OR “Mauritius” [Title/abs tract/keyword] OR “Seychelles” [Title/abstract/keyword] OR “Sudan” [Title/abstract/k eyword] OR “Comoros”[Title/abstract/keyword] OR “Djibout” [Title/abstract/keyword ] OR “Mozambique” [Title/abstract/keyword] OR “Zimbabwe” [Title/abstract/keyword] OR “Malawi” [Title/abstract/keyword] OR “Zambia” [Title/abstract/keyword] OR “Mad agasca” [Title/abstract/keyword] OR “Mayotte” [Title/abstract/keyword]

#10=#8 AND #9

**Web of Science**

1#=Topic (Ts) “Neural tube defects” OR “NTDs” OR “Spinal dysraphisms” OR “Spina bifi da occulta” OR “Closed neural tube defect” OR “Meningocele” OR “Myelomeningocele” O R “Anencephaly” OR “Encephalocele” OR “Iniencephaly”

2#=Ts “Long term adverse effects” OR “Persistent vegetative state” OR “Long term effects” OR “Long term impacts” OR “Long term Outcome” OR “Long term sequelae” OR “ Long standing effects” OR “Long standing impacts” OR “Long standing outcome” OR “Lo ng standing sequelae” OR “Permanent effects” OR “Permanent impacts” OR “Permanent outcome” OR “Permanent sequelae”OR “Extended effects” OR “Extended impacts” OR “E xtended outcome” OR “Extended sequelae”

3#=1# AND 2#

4#=Ts “Survival” OR “Mortality” OR "Child Mortality” OR "Infant Mortality" OR “Paralysi s” OR “Lower limb weakness” OR “Bladder dysfunction” OR “Urinary Incontinence” OR “ Urinary bladder neurogenic” OR “Bowel dysfunction” OR “Constipation” OR “Faecal inco ntinence” OR “Hydrocephalus” OR “Special education needs” OR “Disability” OR “Intellec tual disability” OR “Disabled persons” OR “Disabled Children” OR “Impairment” OR “Han dicapped” OR “Cognitive dysfunction” OR “Ataxia” OR “Uncoordinated movements” OR “ Seizure” OR “Small head” OR “Microcephaly” OR “Facial abnormalities” OR “Skull abnor malities” OR “Neurodevelopmental delays” OR “School attendance”

#5=#1 AND #4

#6= #3 OR #5

7#= Ts “Africa Eastern” OR “East Africa” OR “Africa south of the Sahara” OR "Sub Sahara Africa" OR “Africa” OR “Kenya” OR “Tanzania” OR “Uganda” OR “Burundi” OR “Ethiopia” OR “South Sudan” OR “Somali” OR “Rwanda” OR “Eriteria” OR “Mauritius” OR “Seychelle s” OR “Sudan” OR “Commoros” OR “Djibout” OR “Mozambique” OR “Zimbabwe” OR “Mal awi” OR “Zambia” OR “Madagasca” OR “Mayotte”

#8= #6 AND #7

**African Index Medicus**

“Neural tube defects” [Title/abstract/subject] OR NTDs” [Title/abstract/subject] OR “Sp inal dysraphisms” [Title/abstract/subject] OR “Spina bifida occulta” [Title/abstract/ subject] OR “Closed neural tube defect” [Title/abstract/subject] OR “Meningocele” [Title /abstract/subject] OR “Myelomeningocele” [Title/abstract/subject] OR “Anencephaly” [ Title/abstract/subject] OR “Encephalocele” [Title/abstract/subject] OR “Iniencephaly” [ Title/abstract/ subject]

AND

“Longterm adverse effects” [Title/abstract/subject] OR “Persistent vegetative state [Titl e/abstract/subject] OR “Longterm impacts” [Title/abstract/subject] OR “Longterm outc ome” [Title/abstract/Keyword OR “Longterm sequelae” [Title/abstract/subject] OR “lo ngstanding effects” [Title/abstract/subject] OR “Longstanding impacts” [Title/abstract/ subject] OR “Longstanding outcome” [Title/abstract/subject] OR “Longstanding sequela e” [Title/abstract/subject] OR “Permanent effects” [Title/abstract/subject] OR “Perman ent impacts” [Title/abstract/subject] OR “Permanent outcome” [Title/abstract/subject] OR “Permanent sequelae” [Title/abstract/subject] OR “Extended effects” [Title/abstract /subject] OR “Extended impacts” [Title/abstract/subject] OR “Extended outcome” [Title /abstract/ subject] OR “Extended sequelae” [Title/abstract/ subject]

OR

“Surviv*” [Title/abstract/subject] OR “Mortality” [Title/abstract/subject] OR "Child Mor tality" [Title/abstract/subject] OR "Infant Mortality" [Title/abstract/subject] OR “Paraly sis” [Title/abstract/subject] OR “Lower limb weakness” [Title/abstract/subject] OR “Bladder dysfunction” [Title/abstract/subject] OR “Urine Incontinence” [Title/abstract/ subject] OR “Urinary bladder neurogenic” [Title/abstract/subject] OR “Bowel dysfuncti on” [Title/abstract/subject] OR “Constipation” [Title/abstract/subject] OR “Faecal incon tinence” [Title/abstract/subject] OR “Hydrocephalus” [Title/abstract/subject] OR “Spec ial education needs" [Title/abstract/ subject] OR "Disab*" [Title/abstract/subject] OR "Intellectual disability" [Title/abstract/subject] OR "Disabled persons" [Title/abstract/subject] OR "Disabled Children" [Title/abstract/subject] OR "Impairment" [Title/abstract/subject] OR "Handicapped" [Title/abstract/subject] OR "Cognitive dysfunction" [Title/abstract/subject] OR "Ataxia" [Title/abstract/subject] OR "Uncoordinated movements" Title/abstract/subject] OR "Seizure" [Title/abstract/subject] OR "Microcephaly" [Title/abstract/subject] OR "Small head" [Title/abstract/subject] OR "Facial abnormalities" [Title/abstract/subject] OR "Skull abnormalities" [Title/abstract/subject] OR "Neurodevelopmental delays" [Title/abstract/subject] OR "School attendance" [Title/abstract/subject]

AND
"Africa Eastern" [Title/abstract/subject] OR "Africa south of the sahara " [Title/abstract/subject] OR "Africa" [Title/abstract/subject] OR "Kenya" [Title/abstract/subject] OR "Tanzania" [Title/abstract/subject] OR "Uganda" [Title/abstract/subject] OR "Burundi" [Title/abstract/subject] OR "Ethiopia" [Title/abstract/subjec] OR "South Sudan" [Title/abstract/subject] OR "Somali" [Title/abstract/subject] OR "Rwanda" [Title/abstract/subject] OR "Eriteria" [Title/abstract/subject] OR "Mauritius" [Title/abstract/subject] OR "Seychelles" [Title/abstract/subject] OR "Sudan" [Title/abstract/subject] OR "Comoros" [Title/abstract/subject] OR "Djibout" [Title/abstract/subject] OR 'Mozambique" [Title/abstract/subject] OR "Zimbabwe" [Title/abstract/subject] OR "Malawi" [Title/abstract/subject] OR "Zambia" [Title/abstract/subject] OR "Madagasca" [Title/abstract/subject] OR "Mayotte" [Title/abstract/ subject]
